# REM Sleep is Associated with Cognition and Biomarkers Longitudinally in Older Adults Across the Alzheimer Disease Continuum

**DOI:** 10.64898/2026.01.22.26344642

**Authors:** Valentina Pinilla Manriquez, Era M. Laho, Stephanie A. Marvin, Yazmeen Usman, Michael J. Properzi, Kailee A. Palmgren, Yiwen Rao, Wai-Ying Wendy Yau, Gretchen Reynolds, Keith A. Johnson, Rachel Buckley, Dorene Rentz, Rebecca Amariglio, Susan Redline, Shaun Purcell, Reisa A. Sperling, Ina Djonlagic, Jasmeer P. Chhatwal, Stephanie A. Schultz

## Abstract

Sleep disturbances represent a potentially modifiable risk factor for Alzheimer disease (AD). The extent to which changes in rapid eye movement (REM) sleep are related to the accumulation of AD pathology and brain tissue loss is not well understood. In the current study, ninety-four individuals 74 clinically unimpaired [CU] and 20 clinically impaired [CI]) underwent polysomnography (PSG), cognitive assessments, and neuroimaging. Cross-sectional and longitudinal models examined the associations between PSG outcomes of interest (percentage of sleep period spent in REM [%REM] and REM latency) and pre-clinical Alzheimer’s cognitive composite-5 (PACC5) scores, entorhinal tau-positron emission tomography (PET; 18F-flortaucipir), cerebral amyloid-PET (11C-Pittsburgh Compound B; PiB), AD signature cortical thickness, and hippocampal volume, after adjusting for covariates. Across CU and CI individuals, less time spent in REM sleep and prolonged REM latency were associated with poorer cognition after adjusting for age, sex, and years of education. Additionally, these factors were associated with a greater AD pathologic burden after adjusting for age and sex. Longitudinal data, spanning up to 16 years, demonstrated that the rate of change in cognition and AD biomarkers in the time preceding PSG assessment was also strongly associated with REM characteristics. Our findings highlight the potentially bidirectional relationship between the accumulation of AD pathology and the disruption of REM sleep. Future studies are needed to better understand the longitudinal relationship between sleep characteristics, AD progression, and cognitive decline, and to assess the potential of sleep-focused interventions to alter the course of AD clinical, cognitive, and pathological progression.

**One Sentence Summary:** Cognitive and biomarker data are associated with REM sleep characteristics in unimpaired and impaired individuals cross-sectionally and over time.

## INTRODUCTION

Sleep disturbances are frequent in older adults, and changes in sleep architecture are a physiological hallmark of normal aging.(*1*) Sleep becomes shorter, lighter, and more fragmented, and the prevalence of sleep disorders such as obstructive sleep apnea (OSA) increases with age.(*2*) Sleep plays a critical role in memory processing and cognitive performance,(*3–6*) and changes to sleep architecture and the emergence of sleep disturbances are risk factors for cognitive decline and Alzheimer’s disease (AD).(*7–10*)

A bi-directional relationship between sleep features and AD has been proposed based on studies in murine models and human studies.(*11*, *12*) Soluble amyloid-beta and tau in the central nervous system fluctuate diurnally;(*13–15*) increasing during wakefulness and decreasing during sleep, and sleep deprivation in humans has been shown to increase soluble amyloid-beta relative to controls.(*15–17*) Further evidence(*18*) suggests that amyloid-beta clearance is increased during sleep due to increased interstitial fluid flow and glymphatic clearance, primarily during slow wave sleep (SWS). Alternatively, the presence of amyloid-beta(*19*) and tau tangles(*20*, *21*) further exacerbates disruptions in sleep and changes in sleep architecture, including time spent in rapid eye movement (REM) sleep.

The pathological hallmarks of AD,(*22*) namely the accumulation of extracellular amyloid-beta plaques and intracellular neurofibrillary tau-containing tangles, emerge prior to clinical impairment, with the first identifiable changes occurring a decade or more prior to progressive cognitive impairment. The presence of sleep disturbances throughout the preclinical and clinical disease stages of AD may play an important role in the pathological and clinical progression of AD.(*23*, *24*) In this context, variations in sleep architecture and markers of increased sleep fragmentation may represent both potential noninvasive and cost-effective biomarkers for AD, as well as possible targets for interventions to delay the onset of cognitive decline and AD pathology.

Sleep disturbance can be measured via self-report questionnaires(*25*) and sleep logs, as well as objectively measured actigraphy(*26*) and polysomnography (PSG). PSG is considered the gold standard for clinical characterization of sleep macro-architecture features.(*27*) PSG provides a comprehensive assessment of both depth and quantity of sleep, including characterization of time spent in REM sleep and non-REM (NREM) sleep stages including N1, N2, and N3/slow-wave sleep (SWS) phases, as well as physiological characteristics such as heart rate, apnea-hypopnea indices (AHI), and blood-oxygen saturation.(*28*) Accumulating evidence supports the critical role of distinct macro-architecture sleep features in brain health.(*29–31*) For example, in AD, lower NREM SWS sleep is associated with elevated amyloid burden,(*32*) greater tau accumulation in the medial temporal lobe of older adults,(*33*) and lower cortical thickness, independent of amyloid burden,(*34*) indicating unique relationships between SWS and early AD changes.

However, while previous human studies have mainly focused on cross-sectional relationships between NREM sleep and AD-related cognitive and biomarker alterations, REM sleep accounts for a large portion of the latter half of sleep and is critical for memory consolidation.^35^ In addition, REM latency, the time it takes to enter the first REM cycle, can be modulated by lifestyle factors such as medication use, affecting sleep depth and quantity(^35^), (^36^),(^37^). Recent investigations of REM sleep in the context of AD suggest that reduced REM is associated with amyloid burden and grey-matter volumes. However, results are inconsistent across studies and lack longitudinal evaluation. To address these gaps in the literature, the current study specifically examines REM phase-shift and duration characteristics, including the percentage of total sleep time in REM (%REM) and REM latency (REML), in a mixed cohort of clinically unimpaired and impaired older adults who have completed longitudinal cognitive and multimodal neuroimaging procedures.

## RESULTS

### Description of study sample and sleep architecture characteristics

This study included ninety-four adults (mean age 71 ± 9 years old, 61% female) who completed at-home PSG, magnetic resonance (MRI), and amyloid- and tau-positron emission tomography (PET) neuroimaging, and the Mini-Mental State Examination (MMSE), for which most participants completed prior to the first PSG. A subset of participants (n=73) co-enrolled in multiple studies completed the Preclinical Alzheimer Cognitive Composite (PACC5). The sample characteristics are shown in Table 1. The description of inclusion and exclusion criteria and study design is presented in the Methods, and a description of relevant medication usage is presented in Extended Data Table 1.

**Table 1.** Demographics and PSG characteristics of the cross-sectional sample.

| **Variable** | **N = 94<sup>1</sup>** |
| --- | --- |
| **Age (years)** | 71.4 (54.6, 89.7) |
| **Self-Reported Sex** |  |
| Female | 57 (61%) |
| Male | 37 (39%) |
| **Self-Reported Race** |  |
| White | 74 (79%) |
| Black | 12 (13%) |
| Asian or Other | 8 (8.5%) |
| **Education (years)** | 16.5 (9.0, 20.0) |
| **GDS** | 4.7 (0.0, 23.0) |
| **CDR-SB** | 0.6 (0.0, 8.0) |
| **Total sleep time (min)** | 379.4 (184.0, 571.0) |
| **N1 (%)** | 14.1 (2.7, 37.5) |
| **N1 (min)** | 52.6 (13.0, 129.0) |
| **N2 (%)** | 54.5 (28.6, 80.6) |
| **N2 (min)** | 207.1 (78.0, 430.0) |
| **N3 (%)** | 12.3 (0.0, 40.0) |
| **N3 (min)** | 47.1 (0.0, 172.0) |
| **REM (%)** | 19.1 (3.7, 38.6) |
| **REM (min)** | 73.5 (12.0, 192.0) |
| **REM Latency (min)** | 120.9 (9.5, 471.0) |
| **Sleep Efficiency (%)** | 0.8 (0.5, 1.0) |
| **WASO (min)** | 104.0 (13.0, 323.5) |
| **Apnea-Hypopnea Index (AHI)** | 15.6 (0.3, 65.6) |
| **Avg. HR during sleep** | 60.6 (42.0, 89.0) |

<sup>1</sup>Mean (Min, Max); n (%)
Abbreviations: PSG = polysomnogram; REM = rapid eye movement; N1 (%) = percentage of sleep period spent in stage 1 sleep; N2 (%) = percentage of sleep period spent in stage 2 sleep; N3 (%) = percentage of sleep period spent in N3 sleep; REM (%) = percentage of sleep period spent in REM sleep; and WASO = wake after sleep onset.

PSG scoring was performed by a registered sleep technologist (S.A.M.) in accordance with the AASM scoring manual.(*38*) Extracted features included total sleep time (TST), total time in bed (TIB), sleep efficiency, wake after sleep onset (WASO), apnea-hypopnea Index (AHI), average heart rate during sleep, rapid eye movement (REM) latency determined from sleep onset to the first epoch of REM sleep, as well as duration in minutes and percentage of total sleep time spent in non-REM (N1, N2, and N3) and REM sleep stages. Scatter matrix plots of sleep architecture features are presented in Extended Data Fig. 1. As expected, age was positively correlated with the AHI and WASO and negatively correlated with sleep efficiency (Extended Data Fig. 2a-c and Extended Data Table 2). Females spent less time in N1 and more time in N3, had prolonged REML, had a higher average heart rate during sleep, and had lower AHI compared to males (Extended Data Fig. 2d-h and Extended Data Table 3). There were no significant differences of race on sleep measures. See Extended Data Table 4.

### REM characteristics are associated with cognition

NREM features from this cohort have been examined previously (*34*), therefore primary analyses in the current study focused on characteristics of REM timing and duration, specifically REML and %REM. To examine the cross-sectional relationship between REM characteristics and clinical (clinical dementia rating scale; CDR®) and cognitive outcomes (MMSE and PACC5) we fit linear regression models adjusting for age, sex, and education. Individuals who were CI (CDR > 0; n = 20) had lower %REM and prolonged REML compared to CU individuals (CDR = 0; n = 74; Fig. 1a,d and Table 2). In the entire sample (CI and CU; n = 94), higher global cognition, measured with MMSE, was associated with higher %REM, but not REML (Fig. 1b,e and Table 2). A subset of primarily CU individuals (N = 73) completed additional neuropsychological testing, at the same visit MMSE assessment, sensitive to early changes in cognition (PACC5). Analyses revealed that lower %REM and prolonged REML were associated with worse PACC scores (Fig. 1c,f and Table 2).

**Figure 1.**
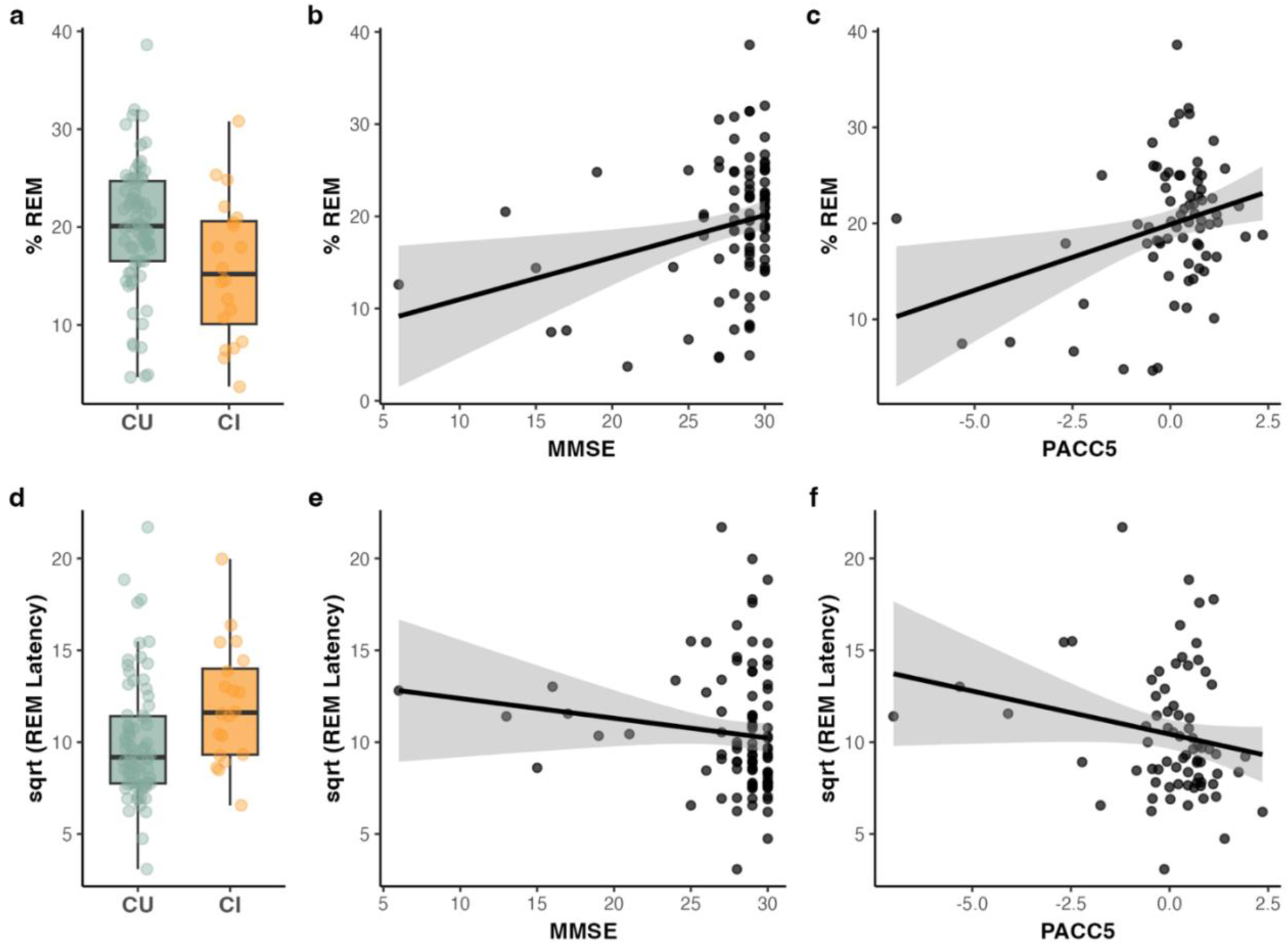
Cognition is associated with REM characteristics. A higher percentage of sleep spent in rapid eye movement stage (%REM) is associated with being cognitively unimpaired (a; CDR = 0), and more favorable scores on the mini-mental state examination (b; MMSE) and preclinical Alzheimer’s cognitive composite 5 (c; PACC5). Prolonged REM Latency was associated with being cognitively impaired (d, CDR > 0), and worse performance on PACC5 (f), but not MMSE (e). Corresponding unstandardized beta-estimates (SE) and p-values from linear regression models, adjusting for age, sex, and years of education, are reported in Supplementary Table 4. Follow-up models examining the associations between REM characteristics and individual cognitive tests spanning domains of working memory, episodic memory, semantic memory, attention, processing speed, and executive function are also presented in Supplementary Table 4.

**Table 2.**
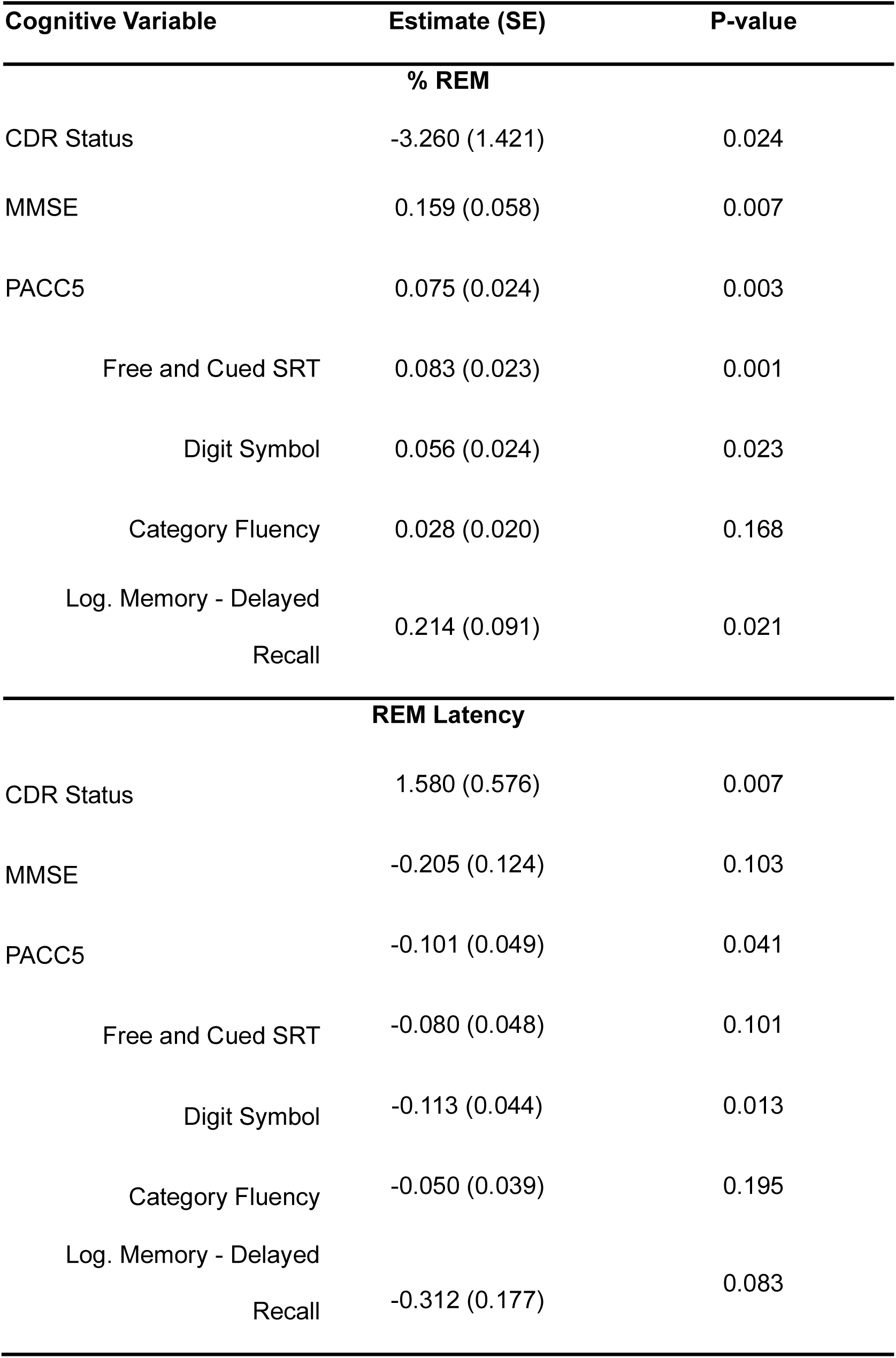
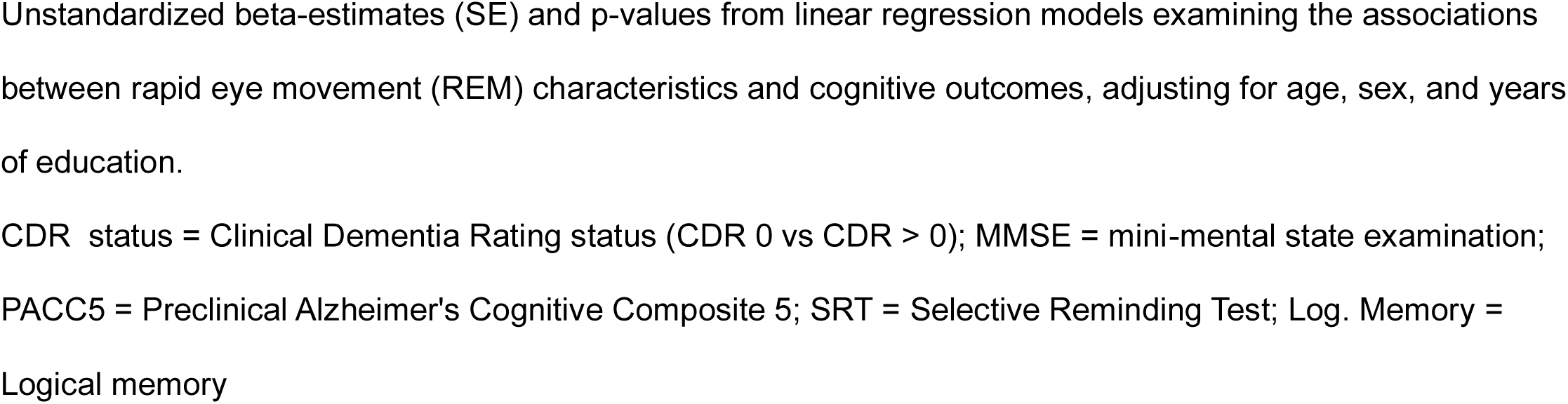
Association between cognition and REM characteristics.

| **Cognitive Variable** | **Estimate (SE)** | **P-value** |
| --- | --- | --- |
| **% REM** |  |  |
| CDR Status | -3.260 (1.421) | 0.024 |
| MMSE | 0.159 (0.058) | 0.007 |
| PACC5 | 0.075 (0.024) | 0.003 |
| Free and Cued SRT | 0.083 (0.023) | 0.001 |
| Digit Symbol | 0.056 (0.024) | 0.023 |
| Category Fluency | 0.028 (0.020) | 0.168 |
| Log. Memory - Delayed<br>Recall | 0.214 (0.091) | 0.021 |
| **REM Latency** |  |  |
| CDR Status | 1.580 (0.576) | 0.007 |
| MMSE | -0.205 (0.124) | 0.103 |
| PACC5 | -0.101 (0.049) | 0.041 |
| Free and Cued SRT | -0.080 (0.048) | 0.101 |
| Digit Symbol | -0.113 (0.044) | 0.013 |
| Category Fluency | -0.050 (0.039) | 0.195 |
| Log. Memory - Delayed<br>Recall | -0.312 (0.177) | 0.083 |

Unstandardized beta-estimates (SE) and p-values from linear regression models examining the associations between rapid eye movement (REM) characteristics and cognitive outcomes, adjusting for age, sex, and years of education.
CDR status = Clinical Dementia Rating status (CDR 0 vs CDR > 0); MMSE = mini-mental state examination; PACC5 = Preclinical Alzheimer's Cognitive Composite 5; SRT = Selective Reminding Test; Log. Memory = Logical memory

Post-hoc analyses, to better understand the relationship between REM characteristics and PACC-5 subtests, were explored in the subset of primarily CU individuals (N = 73) who completed the PACC5. Higher %REM was associated with better scores on all PACC-5 components assessed, except category fluency, while prolonged REML was uniquely associated with poorer performance on the Digit-Symbol Substitution Test. See Table 2.

### Amyloid, tau, and cortical thickness are associated with REM characteristics

Next, we examined whether REM characteristics were associated with cross-sectional *in vivo* cortical (frontal, lateral temporo-parietal and retrosplenial composite (FLR) composite(*39*)) beta-amyloid (11C-Pittsburgh compound B-PET), entorhinal tau (Flortaucipir-PET) levels, hippocampal grey matter volume, and AD signature cortical thickness(*40*). Using linear regression models adjusting for age and sex, we observed that individuals with higher tau, higher beta-amyloid, and lower thickness in an AD-signature composite had lower %REM and prolonged REML (Fig. 2 and Table 3). %REM was not significantly associated with hippocampal volume, though a trend-level effect was seen (p = 0.055, Table 3), and no association between hippocampal volume and REML was observed. To examine potential REM associations with regional cortical or subcortical measures of AD pathology or atrophy, we performed an exploratory analysis using a series of linear regressions for each available cortical and subcortical region (Extended Data Table 5). These exploratory regional analyses revealed widespread associations between REM characteristics and amyloid burden, while relationships between REM and tau and neurodegeneration were more restricted to temporal and parietal regions (Fig. 3 and Extended Data Table 5).

**Figure 2.**
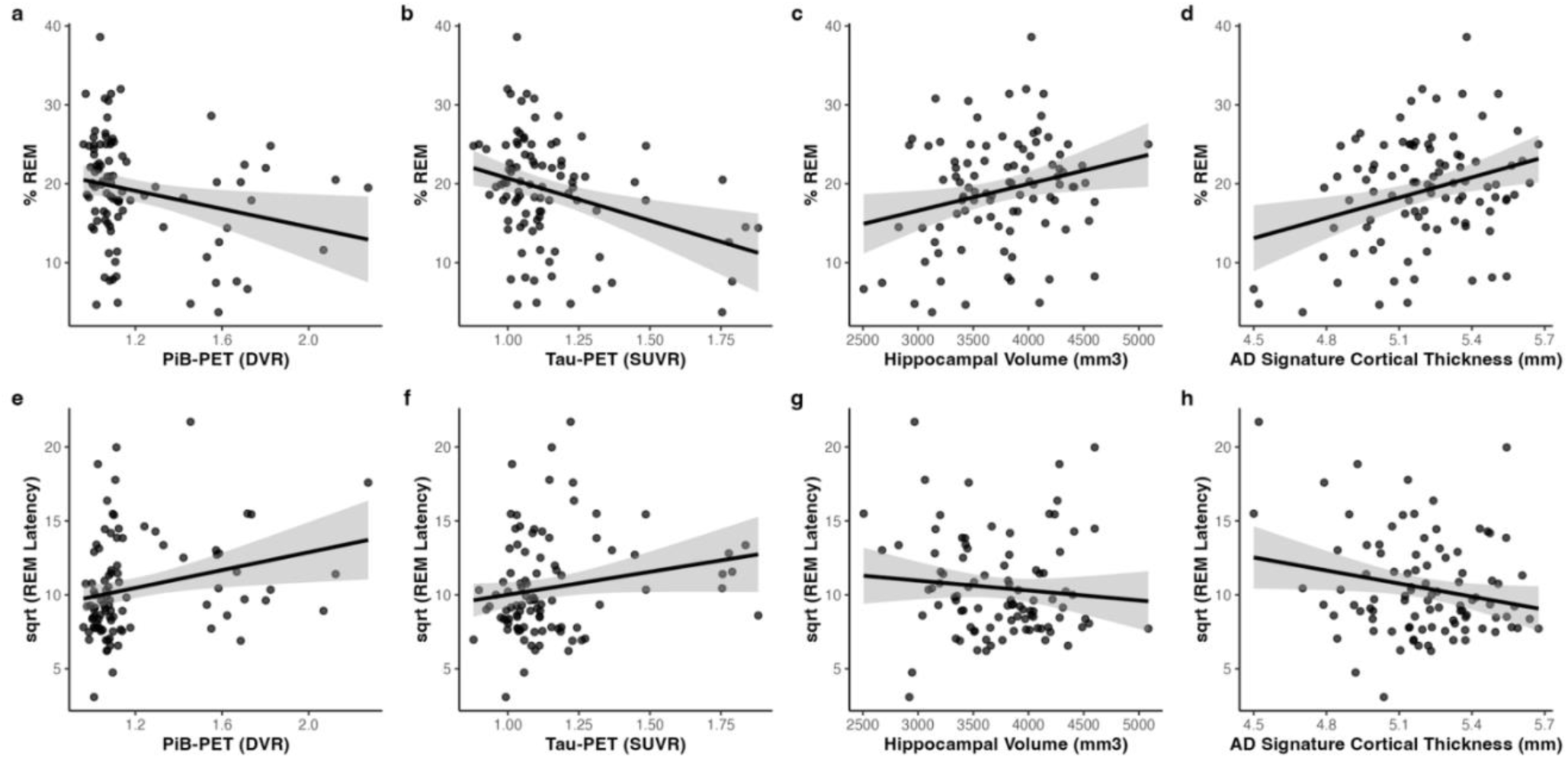
Amyloid, tau, and neurodegeneration are associated with REM characteristics. Individuals who spent less time in rapid eye movement sleep (%REM) had higher amyloid burden (a) and higher tau levels (b), lower hippocampal volume (c) and thinner grey matter (d). Individuals with prolonged REM Latency had higher amyloid (e) and tau (f) burden. There were no observed associations between REM Latency and measures of neurodegeneration (g-h). Corresponding unstandardized beta-estimates (SE) and p-values from linear regression models, adjusting for age and sex, are reported in Supplementary Table 5. Exploratory regional analyses examining the associations between REM characteristics and AD neuroimaging markers from 36 cortical and subcortical regions are presented in Supplementary Figure 3 and Supplementary Table 6.

**Figure 3.**
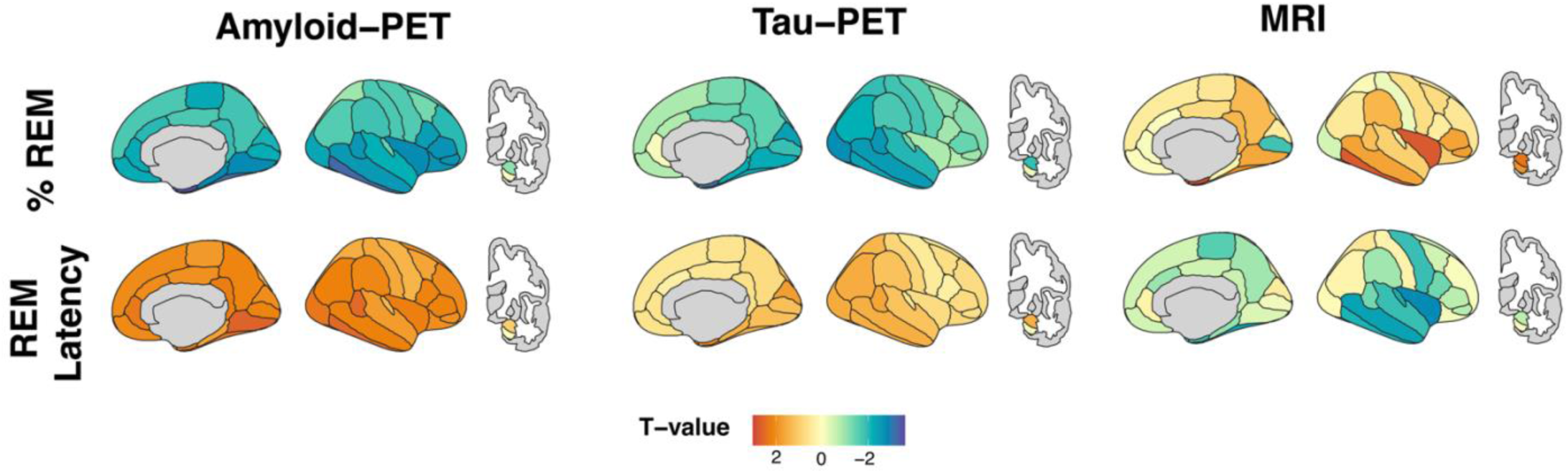
Regional Associations between REM Sleep and AD Biomarkers. T-scores from linear regression models examining the relationship between regional PET and MRI values and %REM or REM Latency, after adjusting for age and sex. There was a negative association between amyloid- and tau-PET values and %REM, and a positive association between MRI (cortical thickness and subcortical volume) and %REM (top row). There was a positive association between amyloid- and tau-PET and REM Latency, and a negative association between MRI and REM Latency (bottom row).

**Table 3.** Association between AD Biomarkers and REM Characteristics.

| **Biomarker Variable** | **Estimate (SE)** | **P-value** |
| --- | --- | --- |
| **% REM** |  |  |
| Cortical composite PiB-PET | -0.009 (0.004) | 0.038 |
| Entorhinal Tau-PET | -0.010 (0.003) | 0.002 |
| ICV-adjusted Hippocampal Vol. | 12.826 (6.608) | 0.055 |
| AD Signature cortical thickness | 0.008 (0.003) | 0.014 |
| **REM Latency** |  |  |
| Cortical composite PiB-PET | 0.019 (0.009) | 0.036 |
| Entorhinal Tau-PET | 0.013 (0.007) | 0.043 |
| ICV-adjusted Hippocampal Vol. | -0.320 (14.059) | 0.982 |
| AD Signature cortical thickness | -0.012 (0.007) | 0.085 |

Unstandardized beta-estimates (SE) and p-values from linear regression models examining the associations
between rapid eye movement (REM) characteristics and AD biomarker outcomes, adjusting for age and sex.
Abbreviations: PiB = Pittsburgh compound B; PET = Positron Emission tomography; ICV = Intracranial volume;
Vol. = volume

We next performed a series of sensitivity analyses to assess the robustness of these associations. As there is evidence to suggest that certain medications may impact sleep architecture (including REM characteristics)(*41*), follow-up analyses were performed to adjust for reported routine usage of SSRIs, SNRIs, benzodiazepines, tricyclic antidepressants, and/or anticholinergic sleep aids (n = 27 reported use of medications of interest, see Extended Data Table 1; note that participants were directed to hold sleep aids for at least 48h prior to PSG). Effect sizes for relationships between REM characteristics and cognitive outcomes remained essentially unchanged after adjusting for medication usages. Adjusting for medication usage generally strengthened the associations between REM characteristics and AD biomarker outcomes. Of note, %REM associations with hippocampal volume and REML associations with AD signature cortical thickness were nominally significant after adjusting for medication usage (Extended Data Table 6).

As increasing age is associated with increased risk of having sleep-disordered breathing, leading to potential changes in overall sleep architecture(*42*), we next re-ran the primary cognitive and biomarker analyses after including AHI as a covariate (Extended Data Table 6). Results were similar following AHI adjustment.

To facilitate harmonization across PET scanners, our main PET analyses were performed with non-partial volume corrected data, due to harmonization techniques across scanners (see Methods for further details). Therefore, we additionally examined the association between REM characteristics and PET measures using partial volume corrected levels in a subset of individuals (n = 36). Observed associations between %REM and amyloid- and tau-PET levels were unchanged. The associations between REML and amyloid- and tau-PET were weakened (Extended Data Table 6).

Lastly, to examine whether relationships between time spent in REM and time spent in other sleep stages were impacting these results, we repeated our primary analyses adjusting for %N1, %N2, and %SWS. Results for %REM remained similar, while findings for REML were weaker after adjusting for %N1 (Extended Data Table 7). Similarly, we repeated our primary analyses with total sleep time (TST) and observed that TST was not associated with any cognitive or AD biomarkers examined (Extended Data Table 8).

### Within-person changes in cognition, amyloid, tau, and neurodegeneration are associated with REM characteristics

To examine whether REM characteristics are a marker of AD progression, we assessed the association between REM measures and the rate of change in cognition and AD biomarkers for those who had completed longitudinal cognitive, MRI, and PET assessments (see Extended Data Table 9). As PSG was added after the start of the study for many participants, a bulk of the longitudinal cognitive and biomarker assessments examined were collected prior to PSG studies (Extended Data Fig. 3). We leveraged these data to examine whether longitudinal changes in cognition, amyloid, tau, and MRI (primarily retrospective with respect to PSG) were associated with REM characteristics. Faster average yearly decline in PACC5 (n = 72; mean follow-up time = 2587 days), mean cortical PiB-PET amyloid accumulation (n = 50; mean follow-up time = 2616 days), PVC-corrected entorhinal tau-PET (n = 65; mean follow-up time = 1696 days), total intracranial volume-adjusted hippocampal volume (HCV) atrophy (n = 92; mean follow-up time = 2038 days), and AD signature cortical thinning (n = 92; mean follow-up time = 2038 days) were associated with lower %REM (Fig. 4 and Table 4). Faster average yearly AD signature cortical thinning was associated with prolonged REML (Fig. 4 and Table 4). Change in MMSE was not associated with REM characteristics (Table 4).

**Figure 4.**
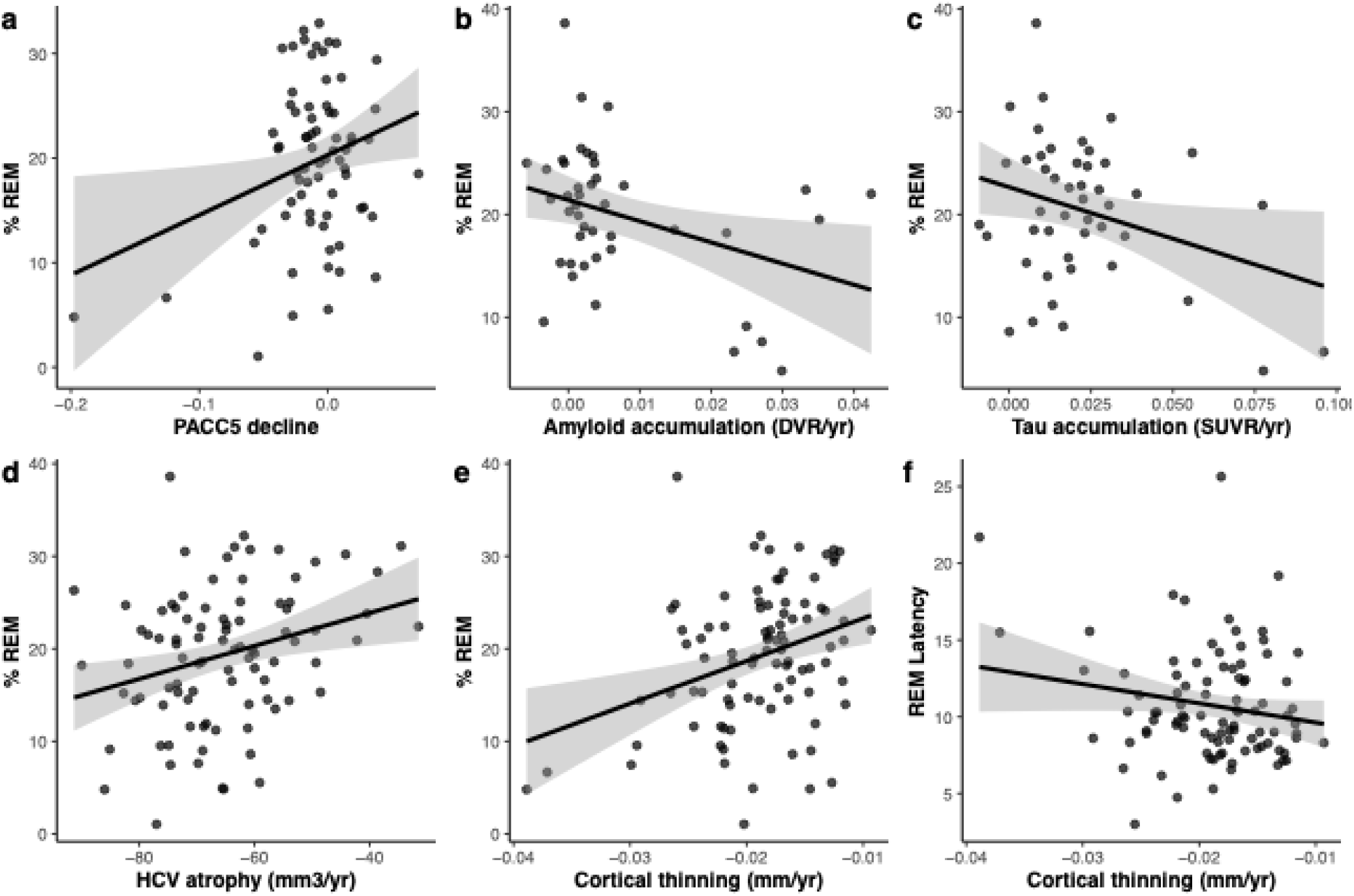
Change in cognition and AD pathology are associated with REM characteristics. Faster average yearly decline in PACC5 (a; n = 72; mean follow-up time = 2587 days), mean cortical PiB-PET amyloid accumulation (b; n = 50; mean follow-up time = 2616 days), PVC-corrected entorhinal tau-PET (c; n = 65; mean follow-up time = 1696 days), total intracranial volume-adjusted hippocampal volume (HCV) atrophy (d; n = 92; mean follow-up time = 2038 days), and AD signature cortical thinning (e; n = 92; mean follow-up time = 2038 days) was associated with lower %REM. Faster average yearly AD signature cortical thinning was associated with prolonged REML (f; n = 92, mean follow-up time = 1305 days) Linear mixed effects models with time-varying cognitive or biomarker measures as the outcome and time*REM characteristics as the main term of interest were fitted with random intercept and slope terms, adjusting for baseline age and sex. Unstandardized beta estimates (standard error) and p-values corresponding to time*REM characteristics are reported in Supplementary Table 10. Models with PACC5 were additionally adjusted for years of education. REM Latency was sqrt-transformed prior to entering into models. Annualized slopes for each outcome were extracted from a linear mixed effects model for plotting purposes.

**Table 2.**
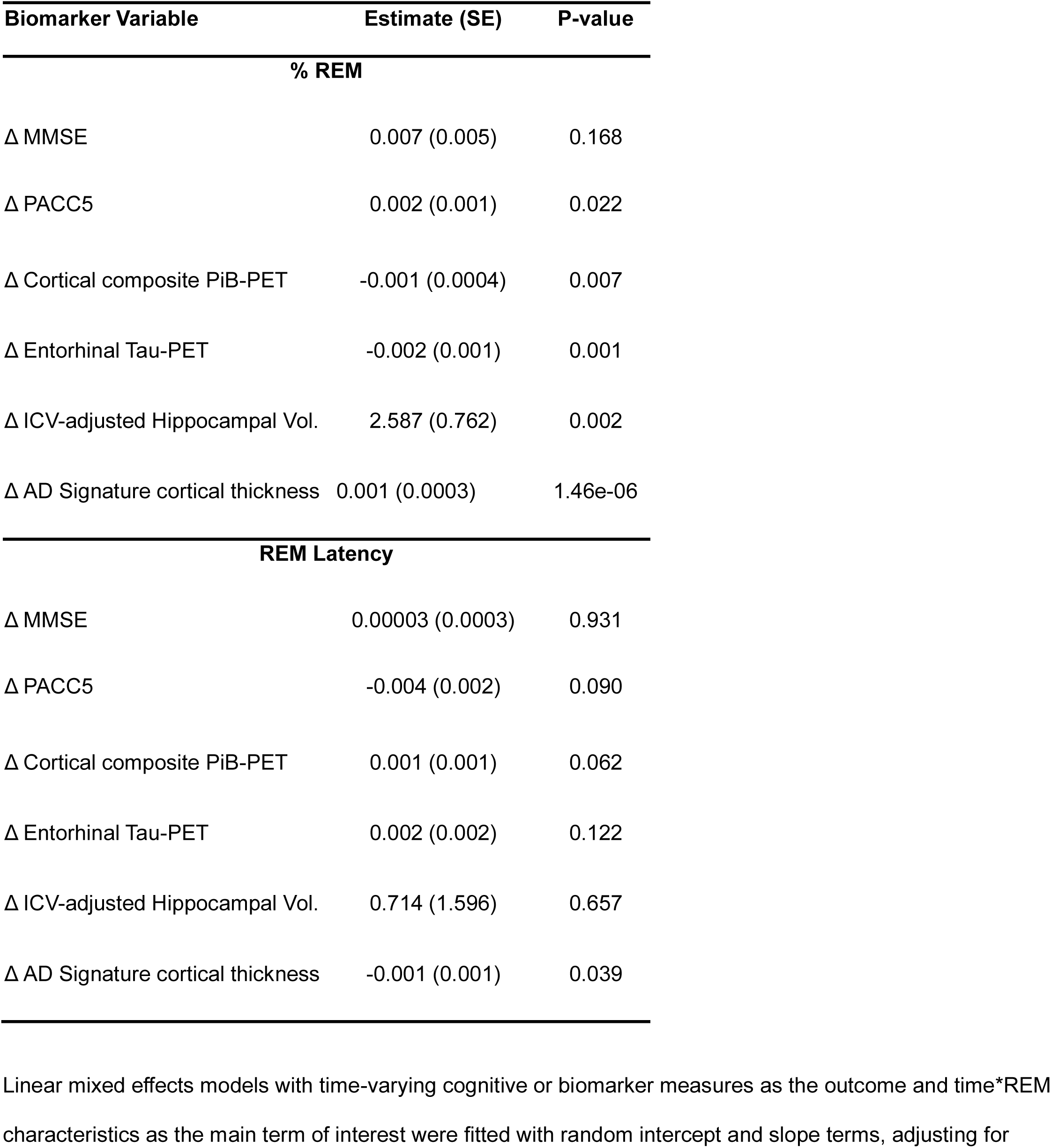
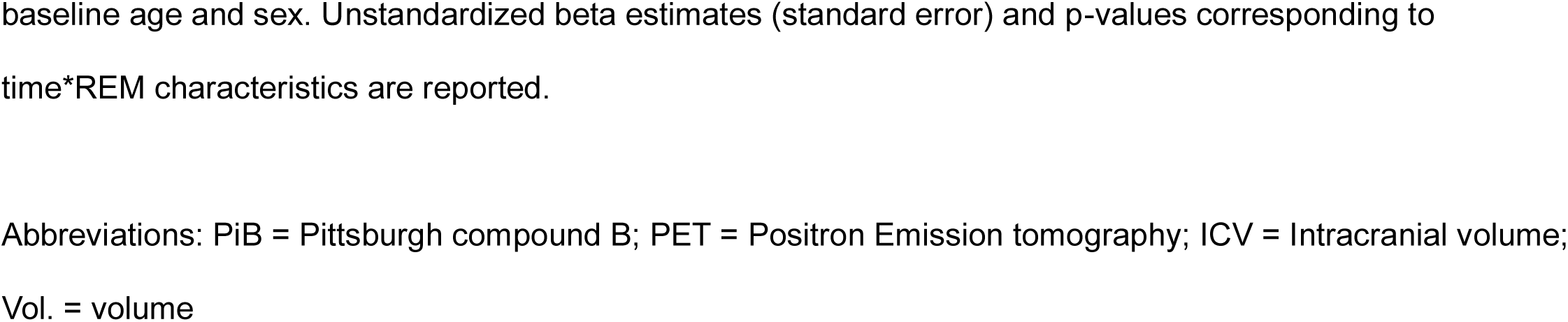
Within-person changes in cognition, amyloid, tau, and neurodegeneration are associated with REM characteristics.

| Biomarker Variable | Estimate (SE) | P-value |
| --- | --- | --- |
| % REM |  |  |
| Δ MMSE | 0.007 (0.005) | 0.168 |
| Δ PACC5 | 0.002 (0.001) | 0.022 |
| Δ Cortical composite PiB-PET | -0.001 (0.0004) | 0.007 |
| Δ Entorhinal Tau-PET | -0.002 (0.001) | 0.001 |
| Δ ICV-adjusted Hippocampal Vol. | 2.587 (0.762) | 0.002 |
| Δ AD Signature cortical thickness | 0.001 (0.0003) | 1.46e-06 |
| REM Latency |  |  |
| Δ MMSE | 0.00003 (0.0003) | 0.931 |
| Δ PACC5 | -0.004 (0.002) | 0.090 |
| Δ Cortical composite PiB-PET | 0.001 (0.001) | 0.062 |
| Δ Entorhinal Tau-PET | 0.002 (0.002) | 0.122 |
| Δ ICV-adjusted Hippocampal Vol. | 0.714 (1.596) | 0.657 |
| Δ AD Signature cortical thickness | -0.001 (0.001) | 0.039 |

Linear mixed effects models with time-varying cognitive or biomarker measures as the outcome and time\*REM characteristics as the main term of interest were fitted with random intercept and slope terms, adjusting for
baseline age and sex. Unstandardized beta estimates (standard error) and p-values corresponding to time\*REM characteristics are reported.
Abbreviations: PiB = Pittsburgh compound B; PET = Positron Emission tomography; ICV = Intracranial volume; Vol. = volume

## DISCUSSION

In a well-characterized cohort of older adults across the AD spectrum, we observed that cognitively impaired individuals spent less time in REM sleep on average and had prolonged REM latency compared to unimpaired individuals. These REM duration and phase-shift characteristics were also associated with global cognition (MMSE), as well as a sensitive presymptomatic cognitive composite (PACC5), both cross-sectionally and longitudinally. These associations remained robust after adjusting for multiple potential confounders, including usage of sleep aids, apnea-hypopnea index, age, sex, and education. Leveraging longitudinal multimodal retrospective neuroimaging, higher amyloid and tau accumulation and neurodegeneration were associated with less REM sleep and prolonged REM Latency. Together, this pattern of results supports a potential bidirectional relationship between REM sleep and AD pathology, and supports the further investigation of sleep disruption as a potentially modifiable risk factor for AD.

There are well-documented examples of physiological changes in sleep architecture across the lifespan, with significant shifts in time spent in N3 and REM sleep apparent by age 60.(*42*) The current study examined age-related sleep differences in a large cognitively unimpaired and impaired cohort ranging from 55 to 90 years old. Our findings support this previous work, reporting decreased sleep efficiency and increased wake after sleep onset with increasing age. Differences in sleep architecture and phase-shift effects were also observed between males and females, with females having lower N1 sleep, higher N3 sleep, prolonged REM latency, increased heart rate during sleep, and lower apnea-hypopnea indices.(*43*) Overall, our results are consistent with the relationships previously observed, including in the Sleep Heart Health Study.(*44*)

Accumulating evidence from mouse and human work suggests that features of sleep architecture are associated with risk of AD dementia and cognitive decline.(*15*, *19*, *33*, *45–48*) SWS plays a critical role in both memory consolidation and fluid dynamics related to Aβ generation and clearance, (*49*, *50*) implicating SWS disruptions in AD pathophysiology. Additionally, emerging evidence suggests REM sleep microstructure is associated with greater neocortical Aβ deposition in cognitively unimpaired older adults,(*51*) and in a recent study focusing on a clinical population including those with MCI and AD, reduced REM sleep and prolonged REM latency was associated with AD biomarkers cross-sectionally,(*45*) but the association between REM characteristics and cognition cross-sectionally and longitudinally remains less understood. Here, we find that cross-sectionally, clinically impaired individuals spent less time in REM sleep and had prolonged REM latency compared to clinically unimpaired individuals. Less time duration in REM and prolonged REM latency were further associated with worse global cognition (MMSE) and lower scores on a sensitive preclinical cognitive composite (PACC). Exploratory analyses revealed that while lower %REM was associated with subcomponents of PACC5, weaker associations were observed with REM latency. With the caveat that the bulk of the available longitudinal cognitive assessments were retrospective (i.e. preceding PSG), we found that greater annualized within-person changes in PACC were associated with lower time spent in REM sleep, but not prolonged REM latency.

Assessing the cross-sectional and longitudinal associations between REM characteristics and multimodal neuroimaging measures, we found that those who spent less time in REM sleep had higher cerebral amyloid and tau PET signal, as well as cortical thinning. Regional analyses revealed widespread associations between REM characteristics and amyloid burden, while relationships between REM and tau and neurodegeneration were more restricted to temporal and parietal regions, observing topographical patterns typically seen in AD. Our longitudinal results strengthened these findings, showing that amyloid, tau, and neurodegeneration accumulation preceding the sleep assessment were linked to poorer REM characteristics. Our results build on previous works linking REM characteristics to white matter health and AD-signature cortical thickness(*52*) (*53*). While mouse models have shown that amyloid can significantly accumulate after just one night of sleep deprivation(*16*), our results show that the longitudinal accumulation of amyloid may influence REM sleep patterns. Similarly, while one study found that REM sleep microarchitecture is associated with amyloid deposition(*51*), this retrospective analysis shows that amyloid may also impact REM in preclinical stages. Furthermore, to the best of our knowledge, this is one of the first studies to examine tau accumulation and REM characteristics using longitudinal measures of tau. As tau begins accumulating in brainstem areas known to play a role in sleep,(*20*) we can infer that tau accumulation greatly impacts REM characteristics in the aging brain. These findings underscore the idea that sleep can be a valuable biomarker of AD long before symptoms arise, and that biomarkers potentially drive REM characteristics.

As %REM may be influenced by time spent in other sleep stages, we assessed the association between REM characteristics and time spent in N1, N2, and SWS stages. We also examine total sleep time as a primary predictor of our cognitive and biomarker outcomes of interest. N1, N2, and SWS did not seem to impact our main findings, and we found no associations between TST and outcomes of interest, suggesting that the associations between REM characteristics and cognition and AD biomarkers are unique. Furthermore, %REM and REM latency measures were moderately correlated in our sample; however, %REM was consistently observed to be more sensitive to the presence and accumulation of cognitive and AD biomarker abnormalities.

This current study utilized multi-modal neuroimaging with both MRI and PET scans. Notably, it included many time points of these imaging markers, which can show pathological changes in the brain that play a role in sleep and REM characteristics. A key feature of our study design also allowed assessment of the reliability of sleep characteristics across our clinically normal and impaired participants. In a subset of individuals who underwent two baseline PSG assessments, there was good reliability in the measures of %N2, average heart rate, and AHI; moderate reliability in sleep efficiency, WASO, %N1, and %REM; and poor reliability in TIB, TST, and REML. Overall, suggesting good reliability in our sample, including cognitively impaired individuals, further points to %REM as a more reliable measure than REM latency.

Additionally, another strength of our study design was the use of at-home sleep measurements, which may provide more ecologically sound data as compared to PSG conducted in an unfamiliar clinic environment. However, several limitations exist. For example, while we are one of the first studies to show longitudinal associations between tau accumulation and sleep characteristics, our analyses rely primarily on retrospective biomarker data, limiting our ability to determine the directional relationship between sleep and biomarkers. Future studies are needed to examine the relationship between sleep architecture and biomarkers using prospective data. Future studies are also needed to determine when changes in sleep occur with respect to the processes of aging and AD pathological change. Such studies are critical to design interventional studies to address sleep disruption as a potentially modifiable risk factor, and to further develop sleep biomarkers relevant to cognitive aging and early pathobiological processes. Lastly, the limitations of available data precluded determining how alcohol use may impact REM sleep, an important consideration in our adult population(*35*).

Older adults commonly report use of at least one prescription medication, some of which can cause drowsiness or interfere with sleep(*43*). In addition, a review and meta-analysis of studies examining the impact of benzodiazepines on dementia risk found that benzodiazepine use increases the risk of dementia(*54*). To address this, in the current study, we investigated whether usage of sleep aids and medications known to impact sleep influenced the observed associations between REM, cognition, and AD biomarkers. After accounting for medication usage, results remained consistent. Cholinesterase inhibitors are also known to affect sleep in AD patients, and the majority of our symptomatic patients were on AD medications. Future studies focused on symptomatic AD patients only should examine AD medication use and dosage and its interactive and synergistic influence on sleep characteristics, cognition, and biomarkers.

Lastly, understanding when in the lifespan AD pathophysiology may be particularly vulnerable to sleep disruption will be critical in determining when such biomarker screening should occur, helping to define time-sensitive windows for developing earlier-life prevention approaches. Although cerebral amyloid accumulation is one of the first key pathological hallmarks of AD and may serve as the instigator of disrupted sleep, other factors may contribute to the severity of sleep problems in patients with AD. In the current study, we find no influence of apnea-hypopnea index on our results, suggesting the presence of sleep apnea does not impact the results presented here. However, older individuals, especially those with other medical conditions, may not have regular physical activity, mealtimes, or lack social interaction, which can impact sleep hygiene(*43*); therefore, they lack strong zeitgebers to entrain their circadian rhythms. Further research is required to tease apart the contributions of these and other psychosocial factors to sleep problems in the AD continuum.

Overall, sleep architecture differed in age, sex, and clinical status. Across CU and CI individuals, REM characteristics were associated with cognitive performance and AD biomarker status. Our findings indicate a novel link between the accumulation of AD pathology and REM patterns, emphasizing a critical need to further understand the longitudinal relationship between sleep characteristics, AD progression, and cognitive decline, particularly in the case of interventions aimed at improving REM sleep quality and how AD (anti-amyloid and anti-tau) therapies impact REM patterns.

## MATERIALS AND METHODS

### Study population

Ninety-four individuals were enrolled in the Sleep Disruption, Tau Accumulation, and Network Dysregulation in Aging, Preclinical, and Symptomatic AD (SONNET) Study. A subset of 50 individuals was recruited through the Harvard Aging Brain Study (HABS)(*55*) or an affiliated HABS substudy.(*56*) The remaining subset of 45 individuals was recruited through memory clinics, community outreach programs, and the Mass General (MGB) Rally website. SONNET, HABS, and HABS-affiliated sub-studies were approved by the MGB Institutional Review Board, and all study participants signed informed consent forms. Participants underwent a battery of cognitive assessments, MRI, and tau- and A*β-*PET imaging, as well as at-home PSG. At study entry, participants were categorized into two arms (clinically normal [N = 74] and Cognitive Concern/Impairment [N = 20]), based on CDR scores (CDR = 0 or CDR > 0, respectively), MMSE scores (MMSE <= 26 and MMSE >15, respectively), and/or a previous diagnoses of mild cognitive impairment or AD. Key exclusion criteria included a history of severe and uncontrolled medical, neurological, and psychiatric conditions, as well as clinically significant sleep-related disorders, including sleep apneas with Apnea-Hypopnea Index (AHI) scores >15, known parasomnias and/or narcolepsy, use of CPAP or supplemental oxygen, history of seizures during sleep, and episodes of excessive daytime somnolence or drowsy driving. Participants taking cholinesterase inhibitors (N = 13) or memantine (N = 2) were included as long as doses had been stable for 8 or more weeks (Extended Data Table 1 for additional details). Although these medications can impact sleep (particularly REM), participants who reported taking selective serotonin-reuptake inhibitors and serotonin/norepinephrine reuptake inhibitors, tricyclic antidepressants, mirtazapine, melatonin, and similar agents were included, if doses were stable for 8 or more weeks. Individuals who took sleep aids (other than melatonin) or benzodiazepines more than twice per week were excluded, while people who took them less than twice per week were included (N = 5) if they were willing and safely able to withhold medication use 48 hours prior to PSG, MRI, or cognitive testing. Individuals who reported currently taking antipsychotics, stimulants, and mood stabilizers were excluded from study enrollment. All participants with available amyloid- and tau-PET, MRI, cognitive assessment, and PSG data were included in the current study.

### Polysomnography

One night of PSG was performed using a Compumedics Somte II device. The montage consisted of six electroencephalogram (EEG) leads (F3, F4, C3, C4, O1, O2, sampling frequency 256 Hz), bilateral electrooculogram (EOG), submental electromyogram (EMG), bilateral anterior tibialis EMG, and standard electrocardiogram (ECG) electrodes. Sensors recommended by the American Academy of Sleep Medicine (AASM) for assessment of sleep-disordered breathing were used, including nasal/oral airflow (thermistor), nasal pressure (Validyne transducer), chest plus abdominal wall motion (inductance plethysmography), and finger pulse oxygen saturation. Participants were encouraged to maintain their habitual evening routines and abstain from sleep aids for the 48 hours preceding the recorded night. PSG scoring was performed by a registered sleep technologist (S.A.M.) in accordance with the AASM scoring manual.(*38*) Main PSG features extracted included total sleep time (TST), total time in bed (TIB), sleep efficiency, wake after sleep onset (WASO, minutes), as well as duration in minutes and percentage of N1, N2, N3, and rapid eye movement (REM) sleep. Arousals were scored according to the AASM manual scoring criteria, which require an abrupt shift of EEG frequency including alpha, theta, and/or frequencies greater than 16 Hz (but not spindles) that last at least 3 seconds with 10 seconds of stable sleep preceding. AHI was calculated as apneas and hypopneas with ≥ 3% oxygen desaturation per hour of sleep. The current study focused on measures of REM including percentage of sleep period spent in REM (%REM) and REM latency (REML).

### Neuropsychological assessment

All participants completed the Clinical Dementia Rating Scale (CDR), Mini-mental State Examination (MMSE) (*57*), Category Fluency, and Boston Naming Test(*58*) as part of their participation in SONNET. A subset of 74 participants additionally completed the Preclinical Alzheimer’s Cognitive Composite-5 (PACC-5) assessment as part of their participation in HABS. The PACC-5 consists of the MMSE, Logical Memory Delayed Story Recall, the Digit-Symbol Substitution Test, the Free and Cued Selective Reminding Test (FCSRT), and FASCAT (*59*). The median (sd) time interval between PSG and neuropsychological assessment for cross-sectional sample was 56 ± 73 days.

### MRI Acquisition and analysis

MRI scans were performed at the Massachusetts General Hospital Athinoula A. Martinos Center for Biomedical Imaging using either a 3T Siemens Trio TIM scanner fitted with a 12-channel head coil or a 3T Siemens MAGNETOM Prisma scanner with a 64-channel head coil. T1-weighted volumetric magnetization-prepared rapid-acquisition gradient-echo (MP-RAGE) images were collected (repetition time/echo time/inversion time=6400/2.8/900 ms, flip angle=8◦, voxel size=1×1×1.2mm3). Image segmentation and regional labeling were performed using FreeSurfer version 6.0. ROIs were adjusted for total intracranial volume. An AD signature cortical thickness measurement ^43^ was created by taking the mean of the thickness (mm) of the following cortical regions from the two hemispheres: middle temporal and inferior temporal, entorhinal, temporal pole, fusiform, superior frontal, rostral and caudal middle frontal, superior and inferior parietal, supramarginal, and precuneus. Bilateral hippocampal volume and AD signature cortical thicknessmeasurements were selected as *a priori* ROIs. Follow-up exploratory regional analyses using examining remaining cortical and subcortical grey-matter volumetrics and thicknesses were also performed. The median (sd) time interval between PSG and MRI for the cross-sectional sample was 1 ± 113 days.

### PET data acquisition and analysis

Amyloid PET was acquired using previously described methods(*55*). In brief, administration of 10 to 15 mCi PiB was followed by a 60 minute acquisition (63 parallel planes: axial field of view: 15.2 cm; in-plane resolution: 4.1 mm full-width at half maximum; slice width: 2.4 mm), using a 39-frame dynamic protocol: 8 × 15s, 4 × 60s, and 27 × 120s. PET data were reconstructed and corrected for scatter, attenuation, and randoms using vendor-supplied software. We calculated the distribution volume ratio (DVR) for PiB-PET based on the Logan graphical analysis technique,(*60*) using the FreeSurfer-defined cerebellar gray reference region of interest (ROI).(*61*) As in our prior publications, PiB retention was summarized using an aggregate cortical ROI including frontal, lateral temporal, and retrosplenial cortices.(*62*) The median time interval between PSG and amyloid PET for cross-sectional sample was 136 ± 315 days.

Tau PET imaging was performed using [^18^F] flortaucipir tau PET tracer using previously described procedures.(*62*, *63*). Regional values for tau PET were calculated based on FreeSurfer-defined cortical ROIs, using cerebellar gray as a reference for calculating standardized uptake value ratios (SUVRs). The entorhinal cortex was selected as our main ROI.(*64*) The median (sd) time interval between PSG and tau PET for cross-sectional sample was 126 ± 237 days. Exploratory analyses examined the broader regional associations across individual FreeSurfer-defined ROIs. Additionally, for PET data acquisition, two cameras were used: Siemens HR+ and GE Discovery. To harmonize the data, images were smoothed with a 3D Gaussian kernel filter with a Full Width at Half Maximum of 6mm prior to processing. Due to harmonization methods, PVC correction could not be applied to harmonized data across scanners. Therefore, sensitivity analyses examining partial volume corrected (PVC) data were performed in the larger subset of participants who scanned on the HR+ scanner. PVC correction was applied using the symmetric geometric transform matrix approach.(*65*)

### Statistical analyses

Analyses were conducted using R version 4.4.2.

#### Dataset creation

For cross-sectional analyses, to minimize the time interval between cognitive, biomarker measures of interest and PSG assessment, data from the PSG and cognitive, MRI, and PET assessments that were completed closest in time to each other were selected for each participant. For longitudinal biomarker analyses, the majority of cognitive, MRI, and PET scans were collected prior to the start of PSG assessments as part of their enrollment in the HABS study. We use this uniquely comprehensive retrospective cognitive and biomarker dataset and selected the PSG assessment closest in time to each participant’s most recent cognitive or biomarker assessment.

Examination for skewness was performed, and a square-root transformation was applied to measures of REM Latency, Avg. HR, Obstructive Apnea index, Hypopnea Index, Central Apnea Index, Total sleep time, and WASO prior to entering into analyses.

#### Description of study sample and sleep architecture characteristics

Univariate linear regression models were used to examine relationships between PSG outcomes and participants’ age and sex, and a one-way ANOVA model was used to examine the relationship between PSG outcomes and participants’ race.

#### Cross-sectional association between REM characteristics and cognition

Multivariate linear regression models adjusting for age, sex, and years of education were used to examine the relationship between REM characteristics (%REM or REML) and CDR status (CDR = 0 vs CDR > 0), MMSE, and PACC-5. Follow-up exploratory analyses further examined the relationship between REM characteristics and individual cognitive tests, including FCSRT, CAT, Digit Symbol Substitution Test, and Logical Memory-Delayed Recall.

#### Cross-sectional association between REM characteristics and AD biomarkers

Primary analyses examining the association between REM characteristics and 1) cortical composite amyloid–PET levels, 2) entorhinal tau-PET levels, 3) hippocampal volume, and 4) AD-signature cortical thickness, were fit with linear regression models adjusting for age and sex. Exploratory regional analyses examined these associations in a large set of 36 cortical and subcortical regions. Model-derived T-values for the association between REM characteristics and amyloid, tau, or grey-matter thickness in each region, after adjusting for age and sex, are depicted on a brain map.

#### Sensitivity Analyses

##### Cross-sectional association between REM characteristics and participant characteristics

To further examine the strength of relationship between %REM and REML and our outcomes of interest, we used AHI as a covariate in the models. Multivariate linear regression models adjusting for age, sex, years of education, and AHI were used to examine the relationship between REM characteristics and participant characteristics. Lastly, medication use was entered into the same model, adjusting for the above-mentioned covariates.

Additionally, total sleep time was entered into the same model to examine its effects on time in REM, adjusting for the above-mentioned covariates.

##### Associations between within-person change in cognition, amyloid, tau, neurodegeneration and REM characteristics

To examine whether REM characteristics can be a marker of AD progression, we assessed the association between REM measures and rate of change in cognition and AD biomarkers in a subset of individuals who had completed longitudinal cognitive, MRI, and PET assessments. Longitudinal cognitive and biomarker assessments were collected mainly before the start of baseline PSG studies. Linear mixed effect models were fit with time-varying PACC-5, cortical composite amyloid–PET levels, entorhinal tau-PET levels, hippocampal volume, or AD-signature cortical thickness as the outcome and a REM characteristic*time interaction as the main term of interest. Models were adjusted for baseline age and sex and include a random intercept and slope term.

## List of Supplementary Materials

Fig. S1 to S4 Table S1 to S9

## Data availability

Biomarker and cognitive data from the Harvard Aging Brain Study (HABS) is available online at http://nmr.mgh.harvard.edu/lab/harvardagingbrain/data. Longitudinal biomarker and cognitive data collected as part of HABS will be made available in periodic data releases at the same web address or via direct request to the corresponding author(s). PSG data can be requested via email addressed to Dr. Jasmeer Chhatwal at. Qualified investigators must abide by the data use agreements, designed to protect the privacy of our participants. Source data are provided with this paper.

## Code availability

Imaging analyses was performed using FreeSurfer v6.0, available at https://surfer.nmr.mgh.harvard.edu/fswiki/DownloadAndlnstall and tools available at https://mrtools.mgh.harvard.edu/index.php?title=Main_Page. As described in the paper, a commonly used, open-source software (R v 4.4.2; Foundation for statistical computing) was used for statistical analyses. Source code available, with publication, at https://github.com/stephaschultz/SONNET_REM_biomarkers.

## Acknowledgements

This research was supported by the NIH (R01AG062667, P01AG036694, K01AG084816, and K23AG084868) and an American Academy of Sleep Medicine Bridge to Success Award (171-BS-17).

## Contributions

All listed authors have substantially contributed to the work presented in this manuscript and have approved the submitted version. Every author agrees to be accountable for their contributions, as well as the contributions of coauthors, to this work, and together they are committed to ensuring a timely resolution for any questions regarding the accuracy or integrity of the included material. Manuscript writing, editing, and the conceptual design were primarily completed by V.P.M., E.M.L., J.P.C., and S.A.S. Dataset creation, data analyses and data visualizations were created by V.P.M. Y.R. M.J.P., and S.A.S. Data collection and processing has been completed by V.P.M., E.M.L., S.A.M., Y.U., M.J.P., K.A.P., W.W.Y., G.R., and S.A.S. The following authors also made contributions to the editing and formatting: K.A.J., S.R., S.P., R.A.S., and I.D

---

**Figure S1.**
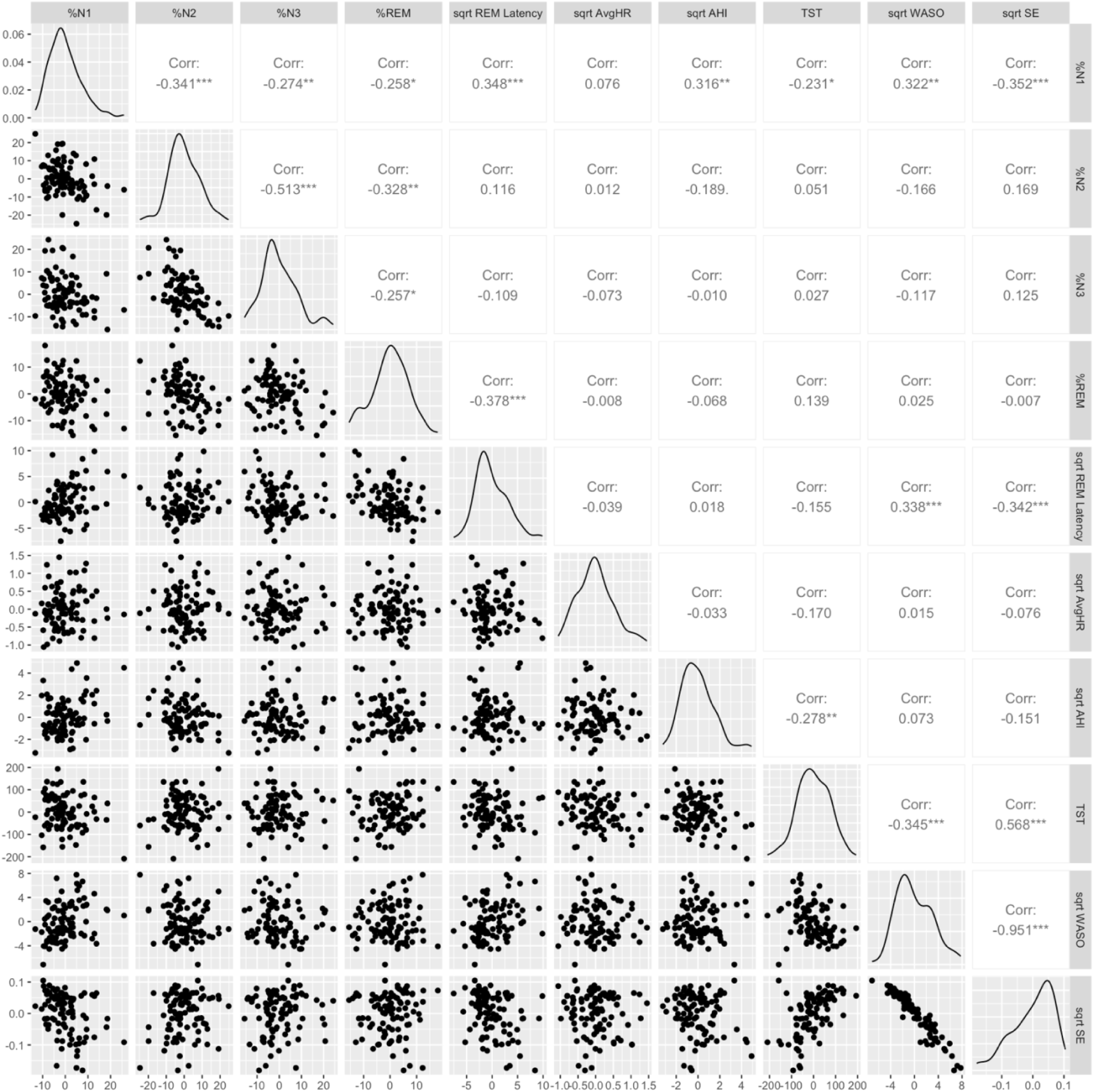
Scatter matrix plots of sleep measures. Correlations were calculated using the Spearman correlation coefficient, where * indicated p < 0.05; ** indicated p < 0.01; and *** indicated p < 0.001. Abbreviations: PSG = polysomnogram; REM = rapid eye movement; N1 (%) = percentage of sleep period spent in stage 1 sleep; N2 (%) = percentage of sleep period spent in stage 2 sleep; N3 (%) = percentage of sleep period spent in N3 sleep; REM (%) = percentage of sleep period spent in REM sleep; and WASO = wake after sleep onset.

**Figure S2.**
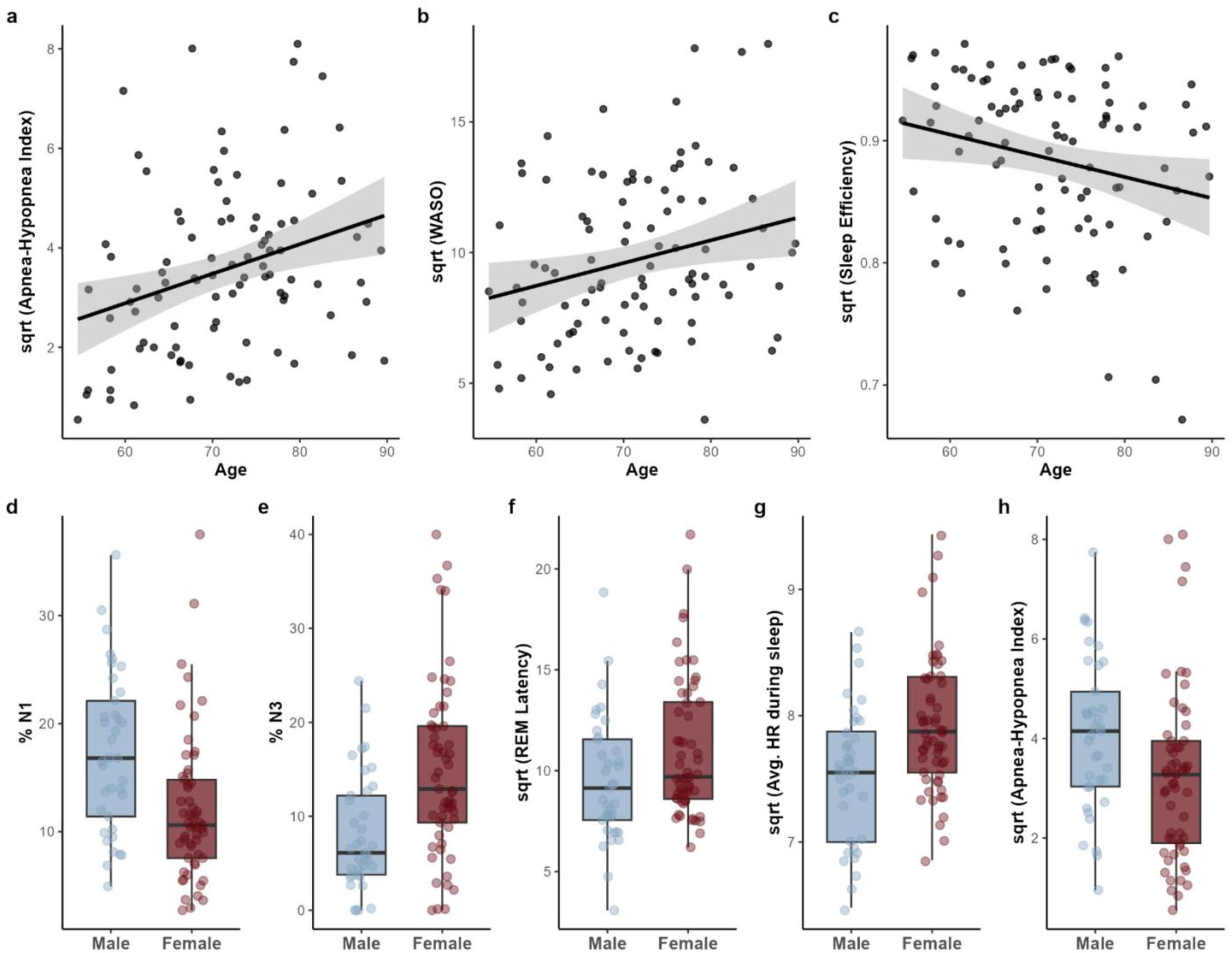
Age and sex are associated with sleep measures. Higher Apnea-Hypopnea Indices (a; AHI), higher Wake After Sleep Onset levels (b; WASO), and lower sleep efficiency (c) are associated with increasing age. Lower percent of sleep time spent in non-rapid eye movement stage 1 (d; %N1), higher percent of sleep spent in non-rapid eye movement stage 3 (e, %N3), prolonged rapid eye movement (REM) Latency (f), higher average heart rate (HR) during sleep (g), and lower AHI (h) are associated with being Female. Corresponding unstandardized beta-estimates (SE) and p-values from linear regression models examining the associations between all sleep measures and age and sex are reported in Supplementary Tables 2-3.

**Figure S3.**
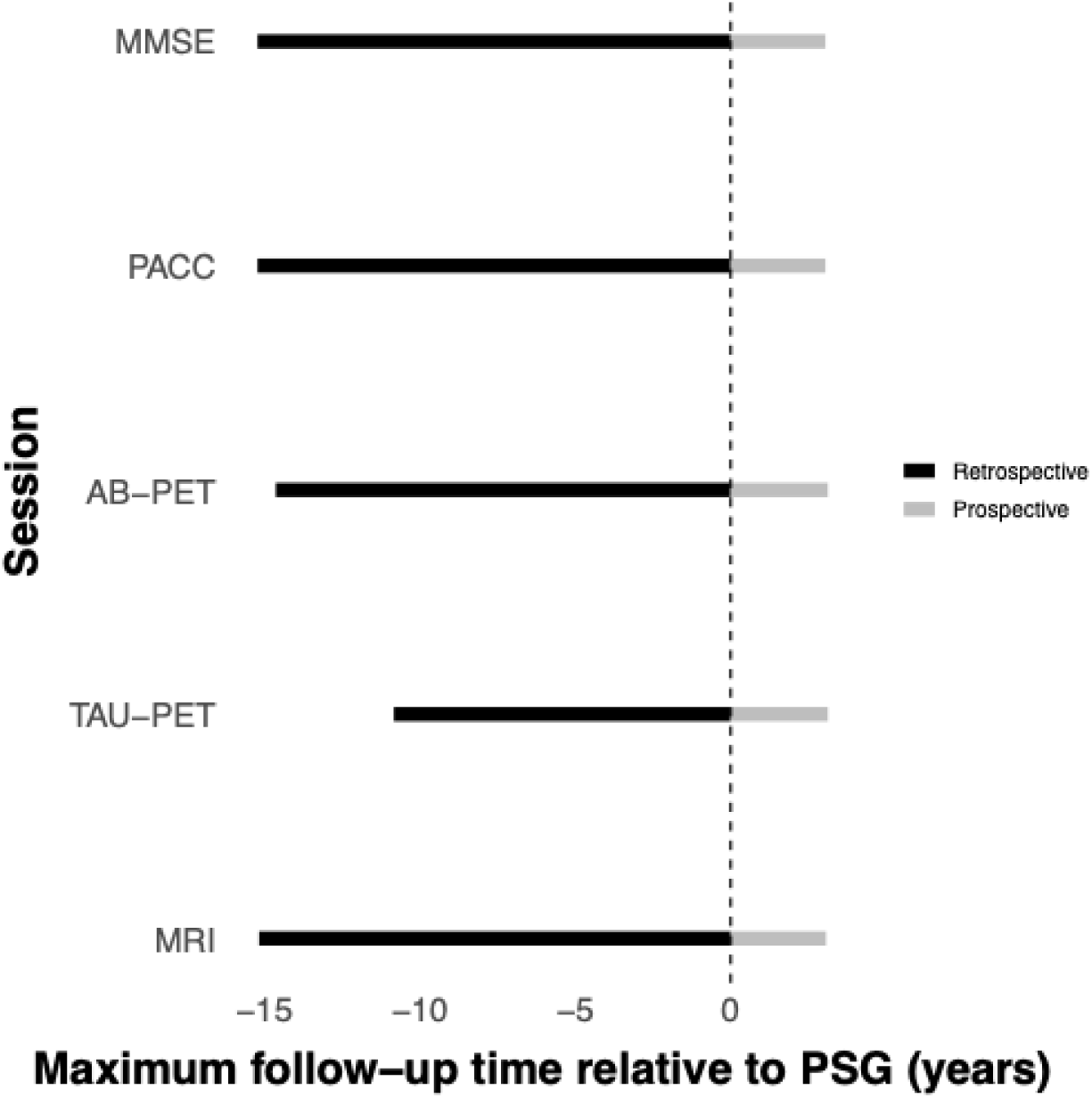
Maximum retrospective and prospective follow-up time for longitudinal cognitive and AD biomarker analyses, relative to PSG.

**Table S1.**
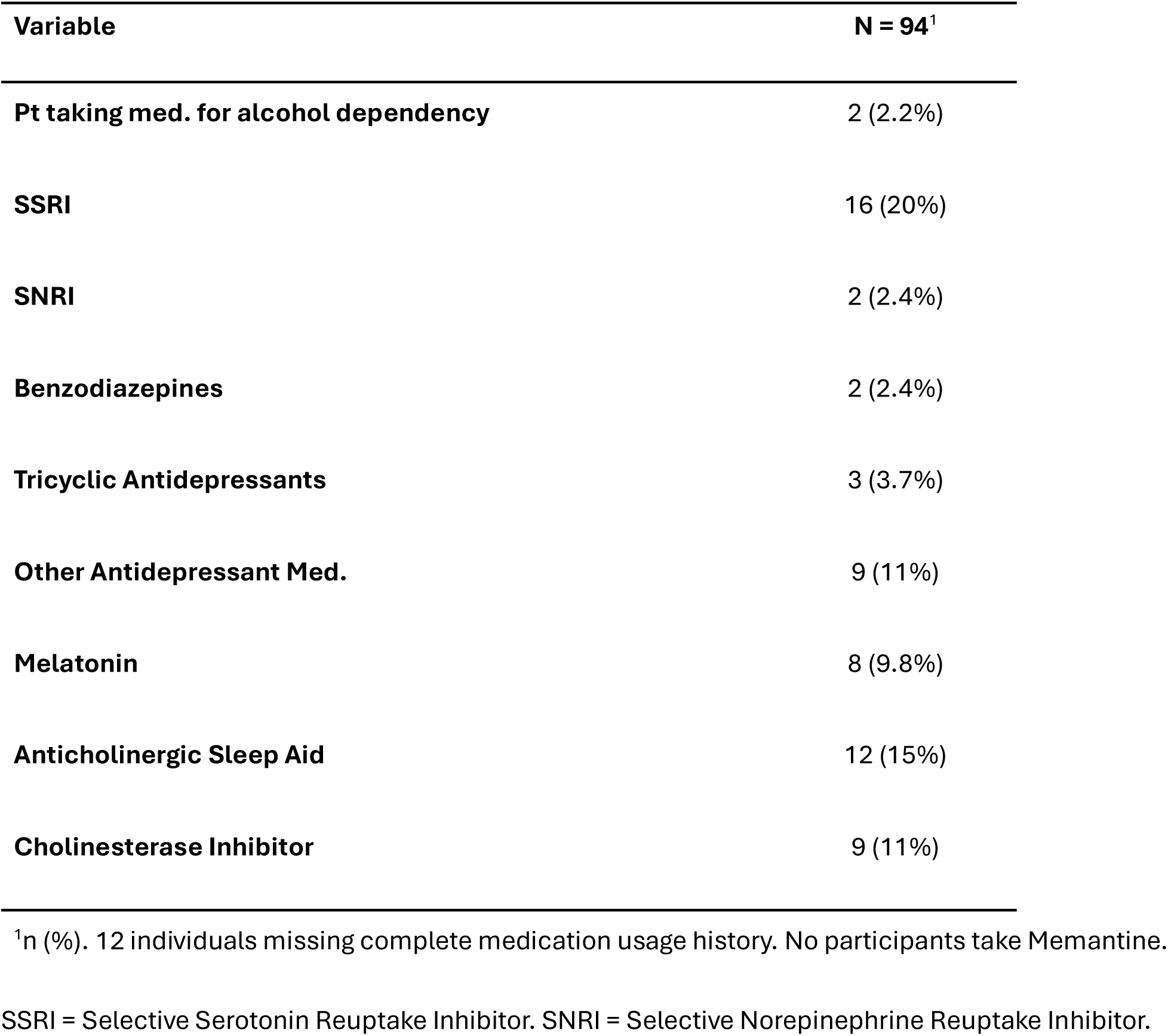
Summary of participant medication usage.

| Variable | N = 94<sup>1</sup> |
| --- | --- |
| Pt taking med. for alcohol dependency | 2 (2.2%) |
| SSRI | 16 (20%) |
| SNRI | 2 (2.4%) |
| Benzodiazepines | 2 (2.4%) |
| Tricyclic Antidepressants | 3 (3.7%) |
| Other Antidepressant Med. | 9 (11%) |
| Melatonin | 8 (9.8%) |
| Anticholinergic Sleep Aid | 12 (15%) |
| Cholinesterase Inhibitor | 9 (11%) |

<sup>1</sup>n (%). 12 individuals missing complete medication usage history. No participants take Memantine.
SSRI = Selective Serotonin Reuptake Inhibitor. SNRI = Selective Norepinephrine Reuptake Inhibitor.

**Table S2.** Effect of age on sleep measures.

| Outcome Variable | Estimate (SE) | P-value |
| --- | --- | --- |
| %N1 | 0.091 (0.088) | 0.306 |
| %N2 | 0.006 (0.106) | 0.957 |
| %N3 | 0.034 (0.106) | 0.751 |
| %REM | -0.131 (0.081) | 0.110 |
| sqrt -REM Latency (min) | 0.052 (0.040) | 0.199 |
| sqrt -Avg. HR during sleep | 0.005 (0.007) | 0.434 |
| sqrt -AHI | 0.059 (0.019) | 0.003 |
| Total sleep time (min.) | -0.946 (0.877) | 0.283 |
| sqrt -WASO | 0.087 (0.036) | 0.017 |
| sqrt -Sleep efficiency | -0.002 (0.001) | 0.028 |

Unstandardized beta-estimates (SE) and p-values from linear regression models examining the associations between sleep measures and age.
%REM = percent of sleep time spent in the rapid eye movement stage; %N1-3 = percent of sleep time spent in non- rapid eye movement stages 1-3; AHI = Apnea-Hypopnea Index; WASO = Wake After Sleep Onset; HR = heart rate.

**Table S3.** Effect of sex on sleep measures.

| Outcome Variable | Estimate (SE) | P-value |
| --- | --- | --- |
| %N1 | -3.702 (1.052) | 0.001 |
| %N2 | -1.886 (1.323) | 0.158 |
| %N3 | 4.880 (1.234) | < 0.001 |
| %REM | 0.703 (1.033) | 0.497 |
| sqrt -REM Latency (min) | 1.085 (0.497) | 0.031 |
| sqrt -Avg. HR during sleep | 0.273 (0.080) | 0.001 |
| sqrt -AHI | -0.592 (0.249) | 0.020 |
| Total sleep time (min.) | 8.092 (11.099) | 0.468 |
| sqrt -WASO | -0.576 (0.463) | 0.217 |
| sqrt -Sleep efficiency | 0.012 (0.010) | 0.220 |

Unstandardized beta-estimates (SE) and p-values from linear regression models examining the associations between sleep measures and sex.
%REM = percent of sleep time spent in the rapid eye movement stage; %N1-3 = percent of sleep time spent in non-rapid eye movement stages 1-3; AHI = Apnea-Hypopnea Index; WASO = Wake After Sleep Onset; HR = heart rate

**Table S4.**
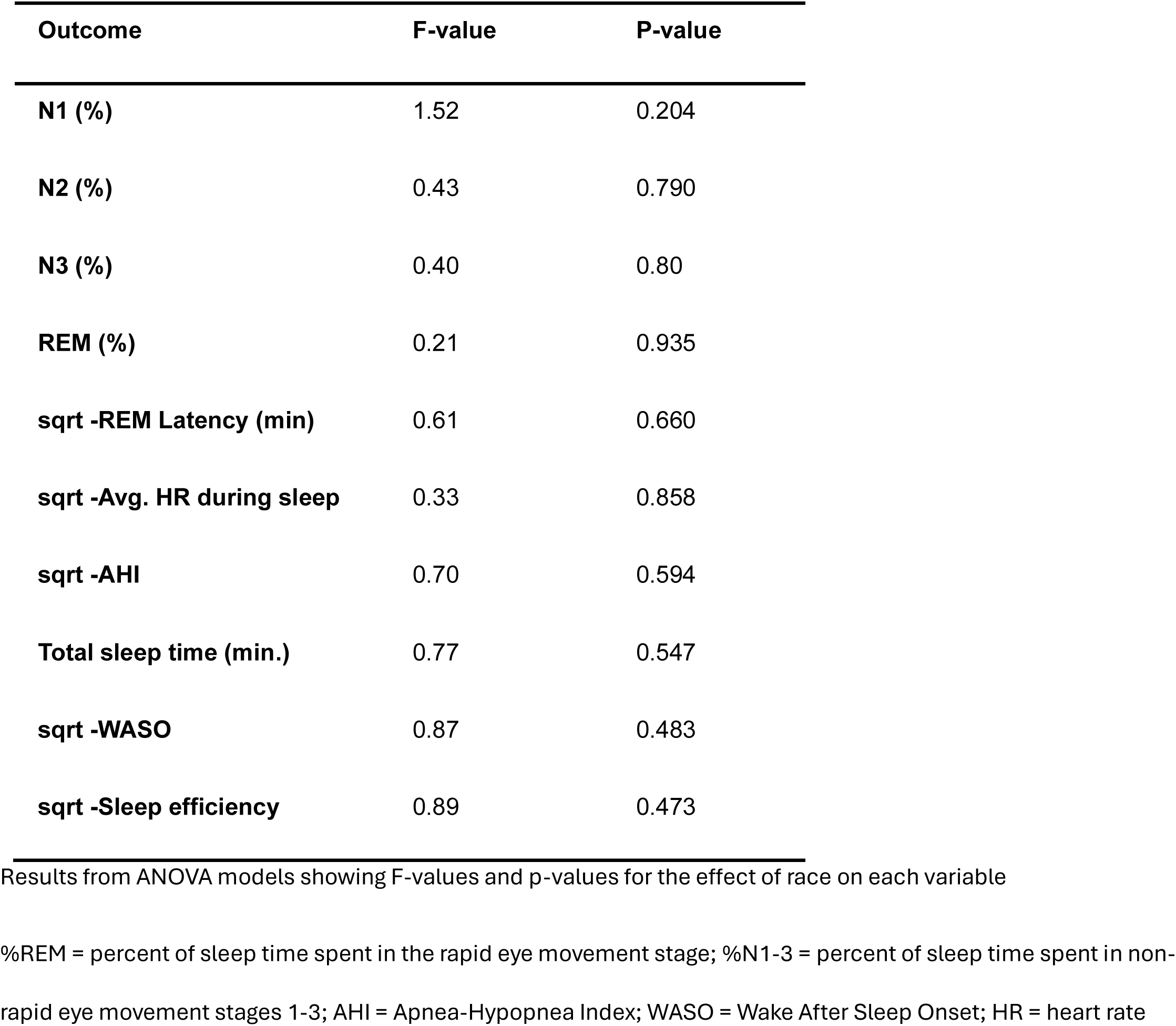
Effect of race on sleep measures.

**Table S5.** Regional associations with REM characteristics.

| region | % REM |  |  |  |  |  | REM Latency |  |  |  |  |  |
| --- | --- | --- | --- | --- | --- | --- | --- | --- | --- | --- | --- | --- |
|  | Amyloid-PET |  | Tau-PET |  | MRI |  | Amyloid-PET |  | Tau-PET |  | MRI |  |
|  | T-Value | P-Value | T-Value | P-Value | T-Value | P-Value | T-Value | P-Value | T-Value | P-Value | T-Value | P-Value |
| amygdala | -1.11 | 0.272 | -1.92 | 0.058 | 2.35 | 0.021 | 0.97 | 0.335 | 1.49 | 0.139 | -0.69 | 0.492 |
| bankssts | -2.56 | 0.012 | -2.57 | 0.012 | 1.23 | 0.223 | 2.49 | 0.014 | 1.65 | 0.103 | -1.32 | 0.190 |
| caudal anterior cingulate | -2.01 | 0.048 | -0.81 | 0.42 | -0.03 | 0.973 | 2.02 | 0.046 | 0.31 | 0.754 | -1.00 | 0.320 |
| caudal middle frontal | -1.31 | 0.194 | -1.10 | 0.276 | 0.76 | 0.447 | 1.65 | 0.103 | 0.72 | 0.474 | -0.50 | 0.622 |
| cuneus | -2.11 | 0.037 | -2.43 | 0.017 | 0.43 | 0.671 | 2.00 | 0.048 | 1.75 | 0.083 | 0.40 | 0.688 |
| entorhinal | -3.56 | 0.001 | -3.16 | 0.002 | 2.90 | 0.005 | 2.24 | 0.028 | 2.05 | 0.043 | -1.86 | 0.066 |
| frontal pole | -2.36 | 0.02 | -2.03 | 0.045 | -0.14 | 0.892 | 1.81 | 0.074 | 1.00 | 0.321 | 1.43 | 0.157 |
| fusiform | -3.20 | 0.002 | -2.33 | 0.022 | 1.98 | 0.051 | 2.4 | 0.018 | 1.44 | 0.154 | -2.4 | 0.019 |
| hippocampus | -0.01 | 0.989 | -0.18 | 0.855 | 1.9 | 0.060 | 0.02 | 0.983 | -0.11 | 0.912 | 0.07 | 0.941 |
| inferior parietal | -1.61 | 0.111 | -2.18 | 0.032 | 0.46 | 0.648 | 2.17 | 0.033 | 1.54 | 0.126 | 0.22 | 0.828 |
| inferior temporal | -3.23 | 0.002 | -2.38 | 0.02 | 2.7 | 0.008 | 2.51 | 0.014 | 1.52 | 0.133 | -2.09 | 0.039 |
| insula | -2.75 | 0.007 | -0.84 | 0.4 | 2.73 | 0.008 | 2.20 | 0.03 | 0.75 | 0.458 | -2.76 | 0.007 |
| isthmus cingulate | -1.92 | 0.058 | -1.56 | 0.122 | 1.37 | 0.174 | 2.22 | 0.029 | 0.66 | 0.513 | -0.57 | 0.573 |
| lateral occipital | -2.36 | 0.021 | -2.73 | 0.008 | -0.46 | 0.644 | 2.32 | 0.023 | 1.56 | 0.123 | 0.07 | 0.944 |
| lateral orbitofrontal | -2.65 | 0.010 | -0.77 | 0.441 | 0.96 | 0.338 | 2.00 | 0.049 | 0.84 | 0.402 | -0.14 | 0.890 |
| lingual | -2.76 | 0.007 | -2.25 | 0.027 | 1.61 | 0.112 | 2.59 | 0.011 | 1.24 | 0.218 | -0.37 | 0.709 |
| medial orbitofrontal | -2.25 | 0.027 | -0.81 | 0.421 | 0.03 | 0.980 | 2.05 | 0.043 | 0.6 | 0.547 | -0.48 | 0.633 |
| middle temporal | -2.54 | 0.013 | -2.51 | 0.014 | 1.67 | 0.098 | 2.16 | 0.033 | 1.5 | 0.137 | -2.4 | 0.018 |
| paracentral | -2.31 | 0.023 | -1.29 | 0.2 | 0.52 | 0.602 | 1.82 | 0.072 | 0.57 | 0.567 | -1.56 | 0.122 |
| parahippocampal | -2.79 | 0.006 | -2.21 | 0.029 | 0.19 | 0.853 | 1.68 | 0.097 | 1.44 | 0.152 | -1.06 | 0.291 |
| pars opercularis | -2.46 | 0.016 | -0.96 | 0.342 | 0.39 | 0.699 | 2.13 | 0.036 | 0.33 | 0.742 | -1.36 | 0.179 |
| pars orbitalis | -2.62 | 0.010 | -0.96 | 0.338 | 1.75 | 0.084 | 1.96 | 0.053 | 0.67 | 0.502 | -0.77 | 0.441 |
| pars triangularis | -2.65 | 0.01 | -1.41 | 0.161 | 1.67 | 0.098 | 2.14 | 0.035 | 0.77 | 0.446 | -0.71 | 0.481 |
| pericalcarine | -2.23 | 0.028 | -1.89 | 0.062 | -1.80 | 0.075 | 2.13 | 0.036 | 1.23 | 0.223 | -0.13 | 0.898 |
| postcentral | -1.70 | 0.092 | -1.81 | 0.074 | -0.19 | 0.851 | 1.61 | 0.111 | 0.87 | 0.386 | 0.22 | 0.824 |
| posterior cingulate | -1.70 | 0.093 | -1.43 | 0.155 | 0.73 | 0.464 | 1.96 | 0.053 | 0.53 | 0.599 | -0.50 | 0.616 |
| precentral | -1.65 | 0.103 | -1.56 | 0.122 | 0.72 | 0.472 | 1.42 | 0.158 | 0.44 | 0.664 | -1.88 | 0.063 |
| precuneus | -1.79 | 0.077 | -1.48 | 0.142 | 1.32 | 0.189 | 2.12 | 0.037 | 0.77 | 0.444 | -0.95 | 0.344 |
| rostral anterior cingulate | -2.27 | 0.025 | -0.08 | 0.935 | -0.07 | 0.941 | 2.29 | 0.024 | 0.59 | 0.558 | 0.24 | 0.811 |
| rostral middle frontal | -2.00 | 0.049 | -1.19 | 0.239 | 0.51 | 0.610 | 2.02 | 0.046 | 0.76 | 0.451 | -0.07 | 0.945 |
| superior frontal | -1.71 | 0.091 | -0.85 | 0.399 | 0.40 | 0.691 | 2.03 | 0.045 | 0.54 | 0.594 | -0.49 | 0.628 |
| superior parietal | -0.98 | 0.332 | -1.92 | 0.057 | 0.10 | 0.918 | 1.52 | 0.131 | 1.39 | 0.169 | 0.48 | 0.636 |
| superior temporal | -2.17 | 0.032 | -2.05 | 0.044 | 0.95 | 0.343 | 1.80 | 0.076 | 1.18 | 0.241 | -1.89 | 0.061 |
| supramarginal | -1.83 | 0.070 | -1.73 | 0.088 | 1.39 | 0.167 | 2.09 | 0.04 | 1.19 | 0.239 | -0.95 | 0.346 |
| temporal pole | -3.00 | 0.004 | -2.53 | 0.013 | 2.79 | 0.006 | 1.66 | 0.100 | 2.16 | 0.034 | -1.62 | 0.110 |
| transverse temporal | -1.42 | 0.158 | -1.36 | 0.176 | 0.37 | 0.712 | 1.33 | 0.187 | 0.53 | 0.597 | -2.13 | 0.036 |

Results from exploratory regional analyses using linear regressions, adjusting for age and sex, examining the relationship between REM or REM Latency and AD pathology (amyloid, tau, and gray matter) in 36 brain regions, T-values and p-values are reported. Significant p-values have been highlighted.

**Table S6.** Sensitivity Analyses for medication usage, AHI, and PCV-PET.

|  | **1. Adjusting for medication usage** |  | **2. Adjusting for AHI** |  | **3. PVC-PET** |  |
| --- | --- | --- | --- | --- | --- | --- |
| **Variable** | **B (SE)** | **P-value** | **B (SE)** | **P-value** | **B (SE)** | **P-value** |
| **% REM** |  |  |  |  |  |  |
| CDR Status | -2.823 (1.409) | 0.049 | -3.272 (1.208) | 0.008 | - | - |
| MMSE | 0.173 (0.068) | 0.013 | 0.154 (0.061) | 0.013 | - | - |
| PACC5 | 0.078 (0.027) | 0.005 | 0.080 (0.025) | 0.003 | - | - |
| Cortical composite PiB-PET | -0.009 (0.004) | 0.053 | -0.009 (0.004) | 0.042 | -0.022 (0.01) | 0.027 |
| Entorhinal Tau-PET | -0.011 (0.003) | 0.002 | -0.010 (0.003) | 0.002 | -0.024 (0.01) | 0.009 |
| ICV-adj. Hippocampal Vol. | 16.817 (7.278) | 0.024 | 13.602 (6.55) | 0.041 | - | - |
| AD Sig. cortical thickness | 0.009 (0.003) | 0.013 | 0.008 (0.003) | 0.012 | - | - |
| **REM Latency** |  |  |  |  |  |  |
| CDR Status | 1.760 (0.636) | 0.007 | 1.579 (0.580) | 0.008 | - | - |
| MMSE | -0.229 (0.147) | 0.124 | -0.205 (0.125) | 0.105 | - | - |
| PACC5 | -0.100 (0.052) | 0.061 | -0.102 (0.049) | 0.041 | - | - |
| Cortical composite PiB-PET | 0.026 (0.010) | 0.008 | 0.019 (0.009) | 0.037 | 0.031 (0.019) | 0.117 |
| Entorhinal Tau-PET | 0.016 (0.008) | 0.044 | 0.013 (0.007) | 0.044 | 0.018 (0.016) | 0.271 |
| ICV-adj. Hippocampal Vol. | -5.952 (16.28) | 0.716 | -0.719 (13.95) | 0.959 | - | - |
| AD Sig. cortical thickness | -0.017 (0.008) | 0.032 | -0.012 (0.007) | 0.083 | - | - |

Unstandardized beta-estimates (SE) and p-values from sensitivity analyses where initial linear regression models examining the associations between rapid eye movement (REM) characteristics and primary cognitive and AD biomarker outcomes were performed after additionally adjusting for 1) medication usage, 2) AHI indices, or 3) using partial volume corrected (PVC) PET values.
CDR status = Clinical Dementia Rating status (CDR 0 vs CDR > 0); MMSE = mini-mental state examination; PACC5 = Preclinical Alzheimer's Cognitive Composite 5; PiB = Pittsburgh compound B; PET = Positron Emission tomography; ICV = Intracranial volume; Vol. = volume

**Table S7.**
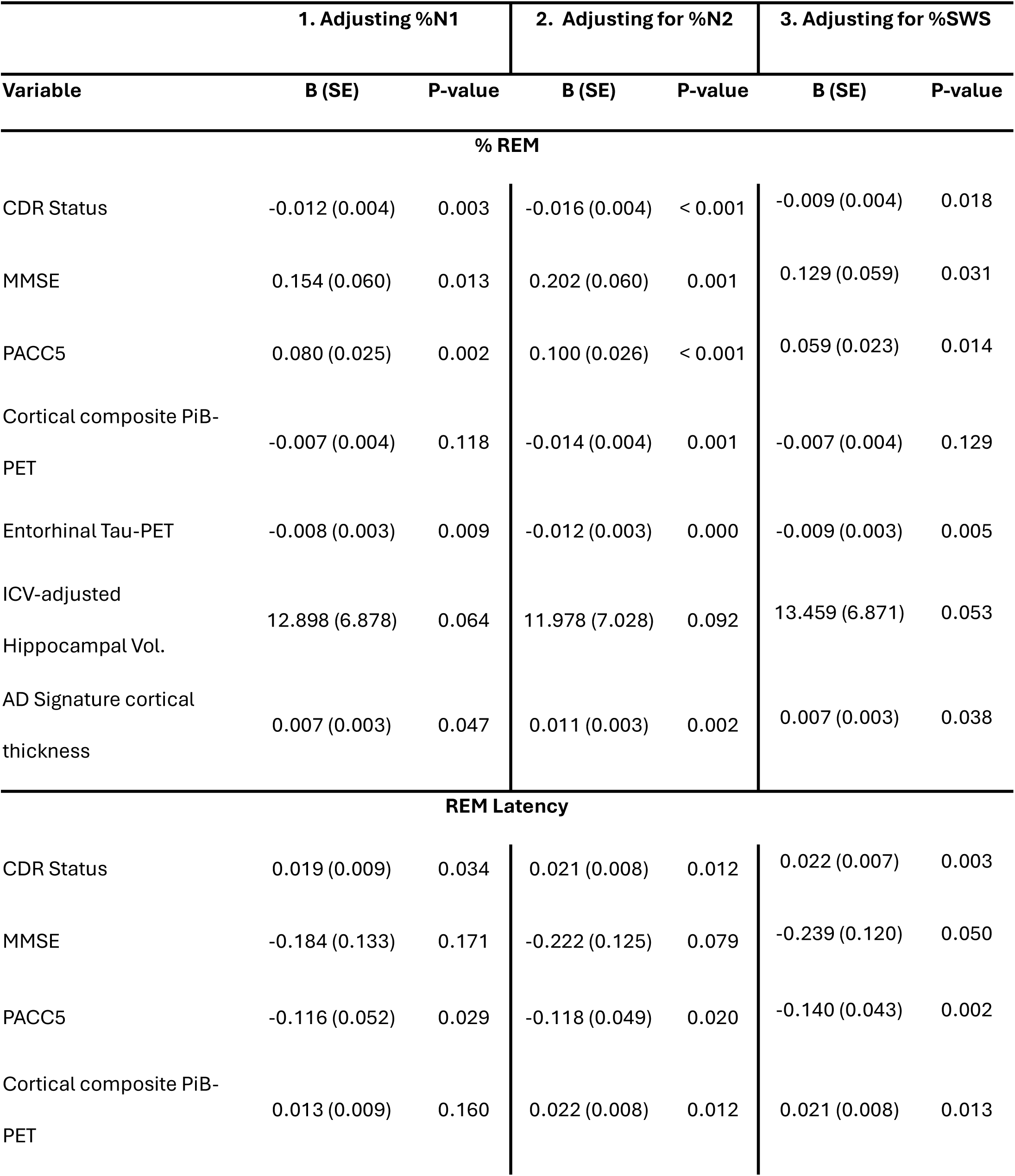
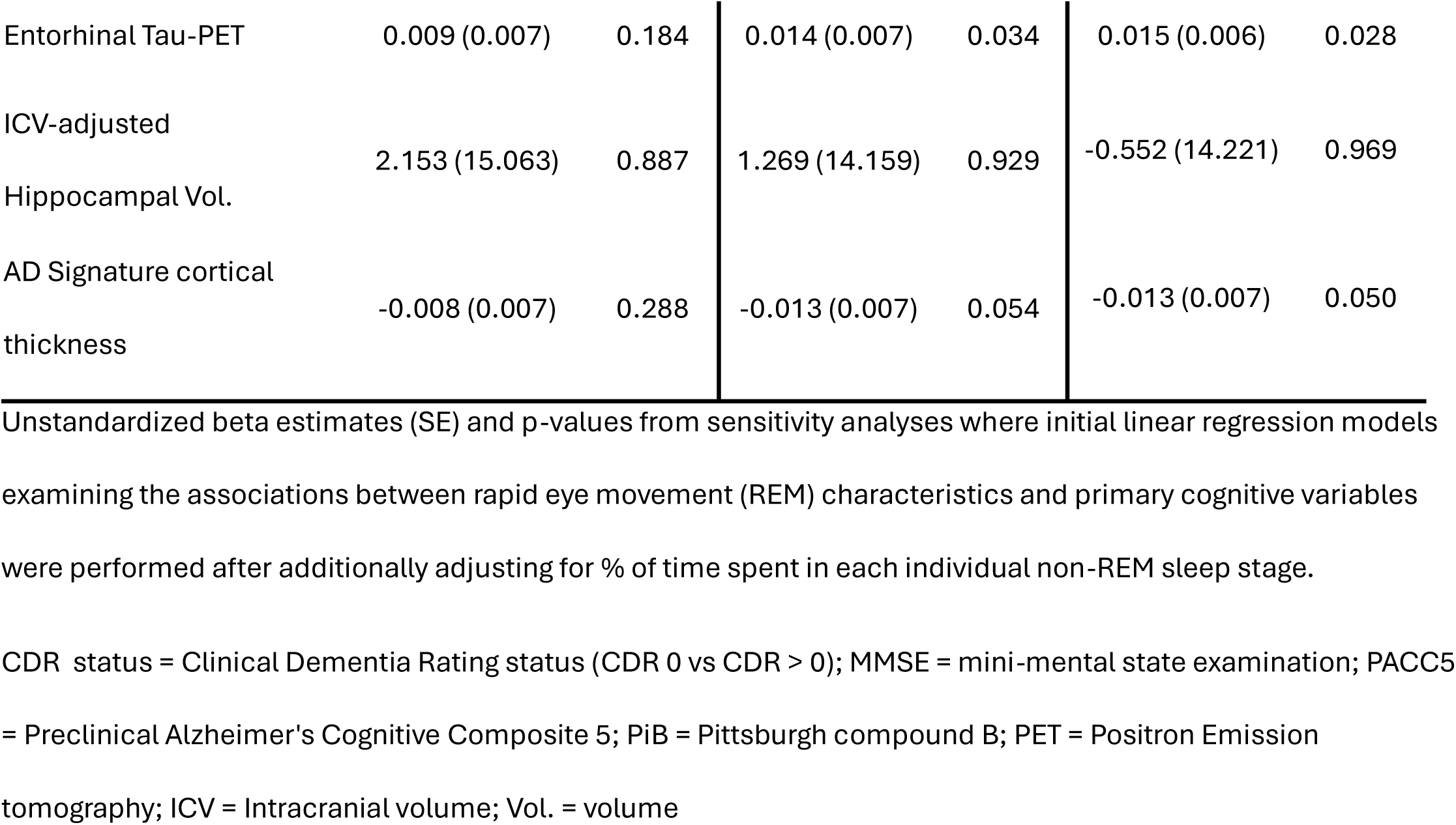
Sensitivity Analyses for %N1, N2, and SWS.

**Table S8.** Sensitivity Analyses for total sleep time.

| Variable | Estimate (SE) | P-value |
| --- | --- | --- |
| CDR Status | 3.623 (13.556) | 0.790 |
| MMSE | -0.010 (0.005) | 0.078 |
| PACC5 | -0.001 (0.002) | 0.611 |
| Cortical composite PiB-PET | 0.001 (0.000) | 0.104 |
| Entorhinal Tau-PET | 0.000 (0.000) | 0.607 |
| ICV-adjusted Hippocampal Vol. | -1.025 (0.613) | 0.098 |
| AD Signature cortical thickness | -0.000 (0.000) | 0.424 |

Unstandardized beta-estimates (SE) and p-values from sensitivity analyses where initial linear regression models were performed to examine associations between total sleep time and primary cognitive and AD biomarker outcomes.
CDR status = Clinical Dementia Rating status (CDR 0 vs CDR > 0); MMSE = mini-mental state examination; PACC5 = Preclinical Alzheimer's Cognitive Composite 5; PiB = Pittsburgh compound B; PET = Positron Emission tomography; ICV = Intracranial volume; Vol. = volume

**Table S9.** Demographics and PSG characteristics of the longitudinal sample.

| **Variable** | **N = 92<sup>1</sup>** |
| --- | --- |
| **Age (years)** | 67.3 (50.3, 89.8) |
| **Self-Reported Sex** |  |
| Female | 57 (62%) |
| Male | 35 (38%) |
| **Self-Reported Race** |  |
| Asian or Other | 6 (6.5%) |
| Black | 11 (12%) |
| White | 75 (82%) |
| **N1 (%)** | 12.3 (1.7, 31.1) |
| **N1 (min)** | 46.7 (8.0, 129.0) |
| **N2 (%)** | 54.5 (23.6, 80.6) |
| **N2 (min)** | 206.5 (45.0, 430.0) |
| **N3 (%)** | 13.9 (0.0, 46.2) |
| **N3 (min)** | 52.2 (0.0, 172.0) |
| **REM (%)** | 19.2 (1.0, 38.6) |
| **REM (min)** | 73.5 (3.0, 165.0) |
| **REM Latency (min)** | 128.4 (9.0, 658.0) |
| **Sleep Efficiency (%)** | 0.8 (0.3, 1.0) |
| **WASO (min)** | 117.3 (4.0, 1,264.5) |
| **Obstructive Apnea Index** | 3.1 (0.0, 24.6) |
| **Central Apnea Index** | 0.6 (0.0, 14.5) |
| **Hypopnea Index** | 12.0 (0.2, 55.4) |
| **Avg. HR during sleep** | 61.2 (43.0, 80.0) |
| **Total sleep time (min)** | 378.0 (191.0, 571.0) |

<sup>1</sup>Mean (Min, Max); n (%)
Abbreviations: PSG = polysomnogram; REM = rapid eye movement; N1(%) = percentage of sleep period spent in stage 1 sleep; N2(%) = percentage of sleep period spent in stage 2 sleep; N3 (%) = percentage of sleep period spent in N3 sleep; REM (%) = percentage of sleep period spent in REM sleep; and WASO = wake after sleep onset.

## References and Notes

1. W. Moraes, R. Piovezan, D. Poyares, L. R. Bittencourt, R. Santos-Silva, S. Tufik, Effects of aging on sleep structure throughout adulthood: a population-based study. Sleep Med. 15, 401–409 (2014).

2. E. Van Cauter, R. Leproult, L. Plat, Age-related changes in slow wave sleep and REM sleep and relationship with growth hormone and cortisol levels in healthy men. JAMA. 284, 861–868 (2000).

3. R. D. Nebes, D. J. Buysse, E. M. Halligan, P. R. Houck, T. H. Monk, Self-reported sleep quality predicts poor cognitive performance in healthy older adults. J. Gerontol. B Psychol. Sci. Soc. Sci. 64, 180–187 (2009).

4. L. Xu, C. Q. Jiang, T. H. Lam, B. Liu, Y. L. Jin, T. Zhu, W. S. Zhang, K. K. Cheng, G. N. Thomas, Short or long sleep duration is associated with memory impairment in older Chinese: the Guangzhou Biobank Cohort Study. Sleep. 34, 575–580 (2011).

5. F. Raven, E. A. Van der Zee, P. Meerlo, R. Havekes, The role of sleep in regulating structural plasticity and synaptic strength: Implications for memory and cognitive function. Sleep Med. Rev. 39, 3–11 (2018).

6. I. Djonlagic, S. Mariani, A. L. Fitzpatrick, V. M. G. T. H. Van Der Klei, D. A. Johnson, A. C. Wood, T. Seeman, H. T. Nguyen, M. J. Prerau, J. A. Luchsinger, J. M. Dzierzewski, S. R. Rapp, G. J. Tranah, K. Yaffe, K. E. Burdick, K. L. Stone, S. Redline, S. M. Purcell, Macro and micro sleep architecture and cognitive performance in older adults. *Nat*. Hum. Behav. 5, 123–145 (2021).

7. S. Bombois, P. Derambure, F. Pasquier, C. Monaca, Sleep disorders in aging and dementia. J. Nutr. Health Aging. 14, 212–217 (2010).

8. C. Liguori, F. Placidi, F. Izzi, M. Spanetta, N. B. Mercuri, A. Di Pucchio, Sleep dysregulation, memory impairment, and CSF biomarkers during different levels of neurocognitive functioning in Alzheimer’s disease course. Alzheimers Res Ther. 12, 5 (2020).

9. B. P. Lucey, J. Wisch, A. H. Boerwinkle, E. C. Landsness, C. D. Toedebusch, J. S. McLeland, O. H. Butt, J. Hassenstab, J. C. Morris, B. M. Ances, D. M. Holtzman, Sleep and longitudinal cognitive performance in preclinical and early symptomatic Alzheimer’s disease. Brain. 144, 2852–2862 (2021).

10. I. Djonlagic, M. Guo, P. Matteis, A. Carusona, R. Stickgold, A. Malhotra, Untreated sleep-disordered breathing: links to aging-related decline in sleep-dependent memory consolidation. PLoS ONE. 9, e85918 (2014).

11. Y. -E. S. Ju, B. P. Lucey, D. M. Holtzman, Sleep and Alzheimer disease pathology--a bidirectional relationship. Nat. Rev. Neurol. 10, 115–119 (2014).

12. C. Wang, D. M. Holtzman, Bidirectional relationship between sleep and Alzheimer’s disease: role of amyloid, tau, and other factors. Neuropsychopharmacology. 45, 104–120 (2020).

13. J. -E. Kang, M. M. Lim, R. J. Bateman, J. J. Lee, L. P. Smyth, J. R. Cirrito, N. Fujiki, S. Nishino, D. M. Holtzman, Amyloid-beta dynamics are regulated by orexin and the sleep-wake cycle. Science. 326, 1005–1007 (2009).

14. Y. Huang, R. Potter, W. Sigurdson, A. Santacruz, S. Shih, Y.-E. Ju, T. Kasten, J. C. Morris, M. Mintun, S. Duntley, R. J. Bateman, Effects of age and amyloid deposition on Aβ dynamics in the human central nervous system. Arch. Neurol. 69, 51–58 (2012).

15. J. K. Holth, S. K. Fritschi, C. Wang, N. P. Pedersen, J. R. Cirrito, T. E. Mahan, M. B. Finn, M. Manis, J. C. Geerling, P. M. Fuller, B. P. Lucey, D. M. Holtzman, The sleep-wake cycle regulates brain interstitial fluid tau in mice and CSF tau in humans. Science. 363, 880–884 (2019).

16. E. Shokri-Kojori, G.-J. Wang, C. E. Wiers, S. B. Demiral, M. Guo, S. W. Kim, E. Lindgren, V. Ramirez, A. Zehra, C. Freeman, G. Miller, P. Manza, T. Srivastava, S. De Santi, D. Tomasi, H. Benveniste, N. D. Volkow, β-Amyloid accumulation in the human brain after one night of sleep deprivation. Proc Natl Acad Sci USA. 115, 4483–4488 (2018).

17. B. P. Lucey, T. J. Hicks, J. S. McLeland, C. D. Toedebusch, J. Boyd, D. L. Elbert, B. W. Patterson, J. Baty, J. C. Morris, V. Ovod, K. G. Mawuenyega, R. J. Bateman, Effect of sleep on overnight cerebrospinal fluid amyloid β kinetics. Ann. Neurol. 83, 197–204 (2018).

18. L. Xie, H. Kang, Q. Xu, M. J. Chen, Y. Liao, M. Thiyagarajan, J. O’Donnell, D. J. Christensen, C. Nicholson, J. J. Iliff, T. Takano, R. Deane, M. Nedergaard, Sleep drives metabolite clearance from the adult brain. Science. 342, 373–377 (2013).

19. J. R. Winer, A. Morehouse, L. Fenton, T. M. Harrison, L. Ayangma, M. Reed, S. Kumar, S. L. Baker, W. J. Jagust, M. P. Walker, Tau and β-Amyloid Burden Predict Actigraphy-Measured and Self-Reported Impairment and Misperception of Human Sleep. J. Neurosci. 41, 7687–7696 (2021).

20. C. H. Lew, C. Petersen, T. C. Neylan, L. T. Grinberg, Tau-driven degeneration of sleep- and wake-regulating neurons in Alzheimer’s disease. Sleep Med. Rev. 60, 101541 (2021).

21. R. E. Irmen, C. M. Carroll, N. J. Constnantino, J. A. Snipes, S. L. Macauley, Tau pathology induced alteration in neuronal activity disrupt sleep and cerebral metabolic rhythms in the P301S PS19 mouse model of tauopathy. Alzheimers Dement. 20 (2024), doi:10.1002/alz.086613.

22. R. A. Sperling, P. S. Aisen, L. A. Beckett, D. A. Bennett, S. Craft, A. M. Fagan, T. Iwatsubo, C. R. Jack, J. Kaye, T. J. Montine, D. C. Park, E. M. Reiman, C. C. Rowe, E. Siemers, Y. Stern, K. Yaffe, M. C. Carrillo, B. Thies, M. Morrison-Bogorad, M. V. Wagster, C. H. Phelps, Toward defining the preclinical stages of Alzheimer’s disease: recommendations from the National Institute on Aging-Alzheimer’s Association workgroups on diagnostic guidelines for Alzheimer’s disease. Alzheimers Dement. 7, 280–292 (2011).

23. C. E. Westerberg, B. A. Mander, S. M. Florczak, S. Weintraub, M.-M. Mesulam, P. C. Zee, K. A. Paller, Concurrent impairments in sleep and memory in amnestic mild cognitive impairment. J. Int. Neuropsychol. Soc. 18, 490–500 (2012).

24. I. Djonlagic, D. Aeschbach, S. L. Harrison, D. Dean, K. Yaffe, S. Ancoli-Israel, K. Stone, S. Redline, Associations between quantitative sleep EEG and subsequent cognitive decline in older women. J. Sleep Res. 28, e12666 (2019).

25. D. J. Buysee, C. F. Reynolds, T. H. Monk, S. R. Berman, D. J. Kupfer, Pittsburgh Sleep Quality Index. American Psychological Association (APA) (2016), doi:10.1037/t05178-000.

26. D. Saleh, S. M. Bertisch, M. Reid, A. Lim, S. Purcell, S. Redline, Actigraphy-derived sleep fragmentation index: convergent validity and associations with clinical outcomes. J. Clin. Sleep Med. 21, 1557–1565 (2025).

27. B. P. Lucey, It’s complicated: The relationship between sleep and Alzheimer’s disease in humans. Neurobiol. Dis. 144, 105031 (2020).

28. J. Feriante, J. F. Araujo, in StatPearls (StatPearls Publishing, Treasure Island (FL), 2025).

29. M. P. Walker, The role of slow wave sleep in memory processing. J. Clin. Sleep Med. 5, S20–6 (2009).

30. Y. F. Lee, D. Gerashchenko, I. Timofeev, B. J. Bacskai, K. V. Kastanenka, Slow wave sleep is a promising intervention target for alzheimer’s disease. Front. Neurosci. 14, 705 (2020).

31. R. F. Gottesman, P. L. Lutsey, H. Benveniste, D. L. Brown, K. M. Full, J.-M. Lee, R. S. Osorio, M. P. Pase, N. S. Redeker, S. Redline, A. P. Spira, American Heart Association Stroke Council; Council on Cardiovascular and Stroke Nursing; and Council on Hypertension, Impact of sleep disorders and disturbed sleep on brain health: A scientific statement from the american heart association. Stroke. 55, e61–e76 (2024).

32. B. A. Mander, S. M. Marks, J. W. Vogel, V. Rao, B. Lu, J. M. Saletin, S. Ancoli-Israel, W. J. Jagust, M. P. Walker, β-amyloid disrupts human NREM slow waves and related hippocampus-dependent memory consolidation. Nat. Neurosci. 18, 1051–1057 (2015).

33. J. R. Winer, B. A. Mander, R. F. Helfrich, A. Maass, T. M. Harrison, S. L. Baker, R. T. Knight, W. J. Jagust, M. P. Walker, Sleep as a Potential Biomarker of Tau and β-Amyloid Burden in the Human Brain. J. Neurosci. 39, 6315–6324 (2019).

34. L. Stankeviciute, J. P. Chhatwal, R. Levin, V. Pinilla, A. P. Schultz, S. Redline, K. A. Johnson, R. A. Sperling, N. Kozhemiako, S. Purcell, I. Djonlagic, Amyloid beta-independent sleep markers associated with early regional tau burden and cortical thinning. Alzheimers Dement (Amst). 16, e12616 (2024).

35. C. Gardiner, J. Weakley, L. M. Burke, G. D. Roach, C. Sargent, N. Maniar, M. Huynh, D. J. Miller, A. Townshend, S. L. Halson, The effect of alcohol on subsequent sleep in healthy adults: A systematic review and meta-analysis. Sleep Med. Rev. 80, 102030 (2025).

36. P. P. Ujma, R. Bódizs, Sleep alterations as a function of 88 health indicators. BMC Med. 22, 134 (2024).

37. E. Ghossoub, L. Geagea, F. Kobeissy, F. Talih, Comparative effects of psychotropic medications on sleep architecture: a retrospective review of diagnostic polysomnography sleep parameters. Sleep Sci. 14, 236–244 (2021).

38. Raman Malhotra, The AASM Manual for the Scoring of Sleep and Associated Events (2023).

39. K. A. Johnson, M. Gregas, J. A. Becker, C. Kinnecom, D. H. Salat, E. K. Moran, E. E. Smith, J. Rosand, D. M. Rentz, W. E. Klunk, C. A. Mathis, J. C. Price, S. T. Dekosky, A. J. Fischman, S. M. Greenberg, Imaging of amyloid burden and distribution in cerebral amyloid angiopathy. Ann. Neurol. 62, 229–234 (2007).

40. B. C. Dickerson, A. Bakkour, D. H. Salat, E. Feczko, J. Pacheco, D. N. Greve, F. Grodstein, C. I. Wright, D. Blacker, H. D. Rosas, R. A. Sperling, A. Atri, J. H. Growdon, B. T. Hyman, J. C. Morris, B. Fischl, R. L. Buckner, The cortical signature of Alzheimer’s disease: regionally specific cortical thinning relates to symptom severity in very mild to mild AD dementia and is detectable in asymptomatic amyloid-positive individuals. Cereb. Cortex. 19, 497–510 (2009).

41. L. J. Klugherz, M. P. Mansukhani, B. P. Kolla, Effects of commonly prescribed medications on sleep: A review of the literature. Mayo Clin. Proc. 100, 856–867 (2025).

42. B. A. Edwards, D. M. O’Driscoll, A. Ali, A. S. Jordan, J. Trinder, A. Malhotra, Aging and sleep: physiology and pathophysiology. Semin. Respir. Crit. Care Med. 31, 618–633 (2010).

43. B. Miner, M. H. Kryger, Sleep in the aging population. Sleep Med. Clin. 12, 31–38 (2017).

44. S. Redline, H. L. Kirchner, S. F. Quan, D. J. Gottlieb, V. Kapur, A. Newman, The effects of age, sex, ethnicity, and sleep-disordered breathing on sleep architecture. Arch. Intern. Med. 164, 406–418 (2004).

45. J. Jin, J. Chen, C. Cavaillès, K. Yaffe, J. Winer, L. Stankeviciute, B. P. Lucey, X. Zhou, S. Gao, D. Peng, Y. Leng, Association of rapid eye movement sleep latency with multimodal biomarkers of Alzheimer’s disease. Alzheimers Dement. 21, e14495 (2025).

46. H. R. Maybrier, J. J. Jackson, C. D. Toedebusch, B. P. Lucey, D. Head, Influence of sleep and cardiovascular health on cognitive trajectories in older adults. Neurobiol. Aging. 152, 34–42 (2025).

47. B. P. Lucey, Sleep alterations and cognitive decline. Semin. Neurol. 45, 333–347 (2025).

48. A. Vaquer-Alicea, J. Yu, H. Liu, B. P. Lucey, Plasma and cerebrospinal fluid proteomic signatures of acutely sleep-deprived humans: an exploratory study. Sleep Adv. 4, zpad047 (2023).

49. B. P. Lucey, A. McCullough, E. C. Landsness, C. D. Toedebusch, J. S. McLeland, A. M. Zaza, A. M. Fagan, L. McCue, C. Xiong, J. C. Morris, T. L. S. Benzinger, D. M. Holtzman, Reduced non-rapid eye movement sleep is associated with tau pathology in early Alzheimer’s disease. Sci. Transl. Med. 11 (2019), doi:10.1126/scitranslmed.aau6550.

50. Y. -E. S. Ju, S. J. Ooms, C. Sutphen, S. L. Macauley, M. A. Zangrilli, G. Jerome, A. M. Fagan, E. Mignot, J. M. Zempel, J. A. H. R. Claassen, D. M. Holtzman, Slow wave sleep disruption increases cerebrospinal fluid amyloid-β levels. Brain. 140, 2104–2111 (2017).

51. C. André, P. Champetier, S. Rehel, E. Kuhn, E. Touron, V. Ourry, B. Landeau, G. Le Du, F. Mézenge, S. Segobin, V. de la Sayette, D. Vivien, G. Chételat, G. Rauchs, Medit-Ageing Research Group, Rapid eye movement sleep, neurodegeneration, and amyloid deposition in aging. Ann. Neurol. 93, 979–990 (2023).

52. M. Altendahl, D. L. Cotter, A. M. Staffaroni, A. Wolf, P. Mumford, Y. Cobigo, K. Casaletto, F. Elahi, L. Ruoff, S. Javed, B. M. Bettcher, E. Fox, M. You, R. Saloner, T. C. Neylan, J. H. Kramer, C. M. Walsh, REM sleep is associated with white matter integrity in cognitively healthy, older adults. PLoS ONE. 15, e0235395 (2020).

53. D. X. Liu, M. N. Braskie, C. Cavaillès, C. Peltz, S. Redline, K. Yaffe, Sleep macro-architecture, nocturnal hypoxemia, and Alzheimer’s disease-related MRI patterns among diverse older adults. Alzheimers Dement. 21, e70280 (2025).

54. E. Simmonds, K. S. Levine, J. Han, H. Iwaki, M. J. Koretsky, N. Kuznetsov, F. Faghri, C. W. Solsberg, A. Schuh, L. Jones, S. Bandres-Ciga, C. Blauwendraat, A. Singleton, V. Escott-Price, H. L. Leonard, M. A. Nalls, Sleep disturbances as risk factors for neurodegeneration later in life. NPJ Dement. 1, 6 (2025).

55. A. Dagley, M. LaPoint, W. Huijbers, T. Hedden, D. G. McLaren, J. P. Chatwal, K. V. Papp, R. E. Amariglio, D. Blacker, D. M. Rentz, K. A. Johnson, R. A. Sperling, A. P. Schultz, Harvard Aging Brain Study: Dataset and accessibility. Neuroimage. 144, 255–258 (2017).

56. C. Gonzalez, K. J. Mimmack, R. E. Amariglio, J. A. Becker, J. P. Chhatwal, C. D. Fitzpatrick, J. R. Gatchel, K. A. Johnson, Z. S. Katz, M. K. Kuppe, J. J. Locascio, O. J. Udeogu, K. V. Papp, P. Premnath, M. J. Properzi, D. M. Rentz, A. P. Schultz, R. A. Sperling, P. Vannini, S. Wang, G. A. Marshall, Associations of the harvard automated phone task and alzheimer’s disease pathology in cognitively normal older adults: preliminary findings. J Alzheimers Dis. 94, 217–226 (2023).

57. M. F. Folstein, S. E. Folstein, P. R. McHugh, “Mini-Mental State”: A practice method for grading the cognitive state of patients for the clinician. J Psychiatr Res (1975).

58. E. Kaplan, H. Goodglass, S. Weintraub, Boston Naming Test. American Psychological Association (APA) (2016), doi:10.1037/t27208-000.

59. K. V. Papp, D. M. Rentz, I. Orlovsky, R. A. Sperling, E. C. Mormino, Optimizing the preclinical Alzheimer’s cognitive composite with semantic processing: The PACC5. Alzheimers Dement (N Y). 3, 668–677 (2017).

60. J. Logan, J. S. Fowler, N. D. Volkow, A. P. Wolf, S. L. Dewey, D. J. Schlyer, R. R. MacGregor, R. Hitzemann, B. Bendriem, S. J. Gatley, Graphical analysis of reversible radioligand binding from time-activity measurements applied to [N-11C-methyl]-(-)-cocaine PET studies in human subjects. J. Cereb. Blood Flow Metab. 10, 740–747 (1990).

61. J. A. Becker, T. Hedden, J. Carmasin, J. Maye, D. M. Rentz, D. Putcha, B. Fischl, D. N. Greve, G. A. Marshall, S. Salloway, D. Marks, R. L. Buckner, R. A. Sperling, K. A. Johnson, Amyloid-β associated cortical thinning in clinically normal elderly. Ann. Neurol. 69, 1032–1042 (2011).

62. K. A. Johnson, A. Schultz, R. A. Betensky, J. A. Becker, J. Sepulcre, D. Rentz, E. Mormino, J. Chhatwal, R. Amariglio, K. Papp, G. Marshall, M. Albers, S. Mauro, L. Pepin, J. Alverio, K. Judge, M. Philiossaint, T. Shoup, D. Yokell, B. Dickerson, T. Gomez-Isla, B. Hyman, N. Vasdev, R. Sperling, Tau positron emission tomographic imaging in aging and early Alzheimer disease. Ann. Neurol. 79, 110–119 (2016).

63. D. T. Chien, A. K. Szardenings, S. Bahri, J. C. Walsh, F. Mu, C. Xia, W. R. Shankle, A. J. Lerner, M.-Y. Su, A. Elizarov, H. C. Kolb, Early clinical PET imaging results with the novel PHF-tau radioligand [F18]-T808. J Alzheimers Dis. 38, 171–184 (2014).

64. C. R. Jack, H. J. Wiste, S. D. Weigand, T. M. Therneau, V. J. Lowe, D. S. Knopman, J. L. Gunter, M. L. Senjem, D. T. Jones, K. Kantarci, M. M. Machulda, M. M. Mielke, R. O. Roberts, P. Vemuri, D. A. Reyes, R. C. Petersen, Defining imaging biomarker cut points for brain aging and Alzheimer’s disease. Alzheimers Dement. 13, 205–216 (2017).

65. O. G. Rousset, Y. Ma, A. C. Evans, Correction for partial volume effects in PET: principle and validation. J. Nucl. Med. 39, 904–911 (1998).

